# Fighting Addictions, improving Lives through COmprehensive rehabilitation with music (FALCO): Protocol for an international randomised controlled trial

**DOI:** 10.64898/2026.02.19.26346573

**Authors:** Monika Geretsegger, Heidi Marie Kirkeng Meling, Alexandra Savinova, Jörg Aßmus, Catherine Lourdes Dy, Trond Stalsberg Mydland, Kenneth Dybdahl, Bjarte Johansen, Stefan Kölsch, Anne Malerbakken, Morten Sommerbakk, Lars Tuastad, Aleksander H. Erga, Jens Hetland, Bianka Karshikoff, Thomas Solgaard Svendsen, Lars Lien, Grethe Emilie Roer, Hans-Andre Røste, Adrian Wangberg, Asena Umay Koçan, Matthew Pelowski, Frank Scharnowski, Giorgia Silani, Milos Stankovic, David Steyrl, Franziska Magel, Rudolf Maisriml, Oliver Scheibenbogen, Julia Fent, Thomas Stegemann, Lucia Gassner, Ingrid Zechmeister-Koss, Tali Gottfried, Moshe Bensimon, Laura Ferreri, Camilla Figini, Laura Fusar-Poli, Pierluigi Politi, Ilona Bidzan-Bluma, Łucja Bieleninik, Judyta Borchet, Daria Makurat, Ilona Poćwierz-Marciniak, Kornelia Kester, Estela Camara, Victor Fernández-Dueñas, Antoni Rodriguez-Fornells, Emma Segura, Adrià Vilà-Balló, Silvia Font-Mayolas, Gloria Brugués, Andrea Maestre Cerdeño, Eva Frigola-Capell, Carlos Schmidt, Daniel Vega, Marcus Herdener, Boris B. Quednow, Saed Belbaisi, Basma Damiri, Christian Gold

## Abstract

**Background:** Substance use disorders (SUD) are associated with a high global burden of disease, with 5.4% of all disability-adjusted life years lost due to alcohol and illicit drugs. Highly prevalent multimorbidity includes polysubstance use, mental health conditions, and other non-communicable and infectious diseases. Where traditional treatments are insufficient alone, music therapy (MT) is highly engaging and improves motivation and reduces craving; however, its long-term effects are unknown. The present study aims to examine long-term effects of active music groups (AMG) and music listening groups (MLG) versus treatment as usual (TAU) on addiction severity, recovery, and other outcomes in people with SUD. Immediate and short-term effects, as well as mechanisms of these interventions, will also be examined.

**Methods:** In individuals with SUD across a wide range of age, gender, socioeconomic, and cultural backgrounds, a parallel 3-arm assessor- and statistician-blinded pragmatic multinational randomised controlled trial (RCT) will determine long-term effects of AMG and MLG versus TAU on addiction severity, recovery, and other outcomes. Patients in participating rehabilitation centres in Europe and the Middle East will be randomised 1:1:1 to AMG plus TAU, MLG plus TAU, or TAU alone in a superiority framework. AMG or MLG will be offered for 1 year from randomisation. The primary outcome will be the composite score of drug/alcohol use of the EuropASI 1 year from randomisation. Embedded studies within a trial (SWATs) will examine (i.) immediate effects of AMG or MLG combined with individual components of TAU combined to determine the best combinations of interventions and (ii.) mechanisms using cognitive testing and brain imaging. With 600 participants randomised, the trial will have 80% power on the primary outcome. Patient representatives, health technology assessment (HTA) bodies, and interventionists have been involved from conception and will ensure feasibility and applicability of the intervention across Europe. The trial is currently recruiting; the first participant was randomised on 26 November 2025.

**Discussion:** This document describes the FALCO RCT, the main part of the FALCO project, which aims to reduce disease burden through innovative, effective, and affordable treatment, and will strengthen research and innovation expertise. Recommendations from FALCO will inform intervention delivery across Europe and beyond, leading to increased safety, effectiveness, and cost-effectiveness, and improved quality of life for individuals with SUD. Stakeholders will be involved in communicating findings across all European countries and regions and ensuring that findings are effectively implemented.

**Trial registration:** ClinicalTrials.gov, NCT07028983, registered 11 June 2025. https://clinicaltrials.gov/study/NCT07028983

## 1. Background

Substance use disorder (SUD) is associated with a high global burden of disease: 4.2% of all disability-adjusted life years lost (DALYs, a combined measure of years of life lost and years lived with disability) are due to alcohol and 1.3% due to illicit drugs*^1^*. Multimorbidity is highly prevalent and includes polysubstance use (using several substances), co-occurring mental health conditions (e.g., depression, anxiety, trauma-related disorders), and various sequelae that could be prevented by more effective SUD treatment, including infectious and non-communicable diseases. SUD is also related to poverty and crime through multiple bidirectional links, increasing the societal importance of improving SUD rehabilitation. SUD impacts “everywhere, everything, everyone” in the EU*^2^*. SUD rates remain high, and many patients do not benefit from or drop out of existing pharmacological and non-pharmacological interventions*^3^.* Notably, approved pharmacological treatment options are not available for many SUDs, and non-pharmacological approaches are still the backbone of SUD treatment*^4^*. Partly due to the limited evidence for SUD interventions, European guidelines differ considerably in their recommendations. In clinical practice, a wide range of psychotropic medications are prescribed for the treatment of co-occurring mental health conditions or behavioural problems associated with SUD*^5^*. Some non-pharmacological interventions, such as motivational interviewing (MI), are recommended widely and have become common practice*^6,7^*, while others, such as mindfulness-based intervention (MBI) or music therapy (MT), are not as widespread yet, but are recommended in some SUD guidelines*^6^*. Arts and culture interventions, in general, are more widely recommended as part of a comprehensive approach to mental health*^8^*.

In MT, defined as the clinical and evidence-based use of music interventions to accomplish individualised goals within a therapeutic relationship by a credentialed professional*^9^*, a music therapist uses music and musical activities to support individuals or groups in reaching specific therapeutic goals. This includes activities such as music listening and active music making, which are tailored to participants’ cultural context and individual needs. While people in all human cultures use music to connect socially and to express themselves, forms of expression vary widely. In people with SUD and other mental health problems, the same type of music can be used in beneficial or harmful ways*^10–12^*, with complex links between music experiences and addiction*^13–16^*. The prevalent links between music and substance use across music genres may be explained through underlying traits of creativity, curiosity, risk-taking and sensation-seeking*^17^*. The complex links between music and addiction may imply that specialised MT expertise is needed to maximise health-promoting uses and minimise harmful uses of music in SUD*^13,18^*. Along with tailoring to specific sub-populations, which may be defined geographically, culturally, ethnically, demographically, or clinically, harmonisation and standardisation of intervention protocols across countries and local cultures are also desirable to ensure replicability of research results and equitable evidence-based provision of care*^19^*. Currently, MT services and training availability vary widely across and within countries (https://emtc-eu.com/).

Putative mechanisms of MT in SUD are related to the brain’s reward system. Music engages the brain’s reward system similarly to addictive substances*^20–22^*. However, conversely to addictive psychotropic substances with addictive potential, it promotes positive reinforcement, emotional regulation, and stress reduction in the short- and long-term. Therefore, music may provide an effective and affordable addition to reduce SUD related burden*^23^*. Music is consistently rated as one of the most intrinsically rewarding activities*^24^*, and music reward recruits neurochemical and neurophysiological pathways that overlap with the reward response elicited by recreational substances, enabling its therapeutic use. Key components of the reward circuit in the brain include the nucleus accumbens, amygdala, prefrontal cortex, hippocampus, and ventral tegmental area*^25,26^*. Similar brain networks are involved in music listening and playing*^22,25^*. Indeed, like reward processing, music can evoke strong emotional reactions, retrieve memories and associations that will prospectively guide future behaviour, and create expectations whose violation or satisfaction is important in learning from experience*^27–30^*. Dopamine is central to reward processing and is involved in the experience the effects of psychotropic substances*^31^* as well as music*^20,31^*. However, chronic SUD leads to an attenuated response in reward regions upon natural reinforcers (discrepancy between expected and actual experience)*^32^*.

In this context, it is notable that MT for people with SUD has shown robust immediate effects on both craving and motivation, outcomes which are related to the anticipatory component of the reward system*^23^*. Reduction of craving (or the ability to control it) and the cognitive motivation to change substance use behaviour are vital in the process of drug rehabilitation. Rigorous evidence synthesis from a Cochrane review of 21 randomised controlled trials (RCTs) suggests that MT reduces craving (Fig. 1a), increases motivation for change (Fig. 1b), and improves short-term mental health outcomes^7^. A more recent RCT, identified in an update search in 2024, provided additional evidence on short-term outcomes (anxiety, depression, craving, emotion regulation) of group MT versus active control*^33^*. No other planned, ongoing, or completed studies were identified in our searches. To establish MT as a promising approach more systematically across Europe, more knowledge is needed on the comparative and long-term effects of different types of MT in the context of other existing treatments.

**Figure 1.**
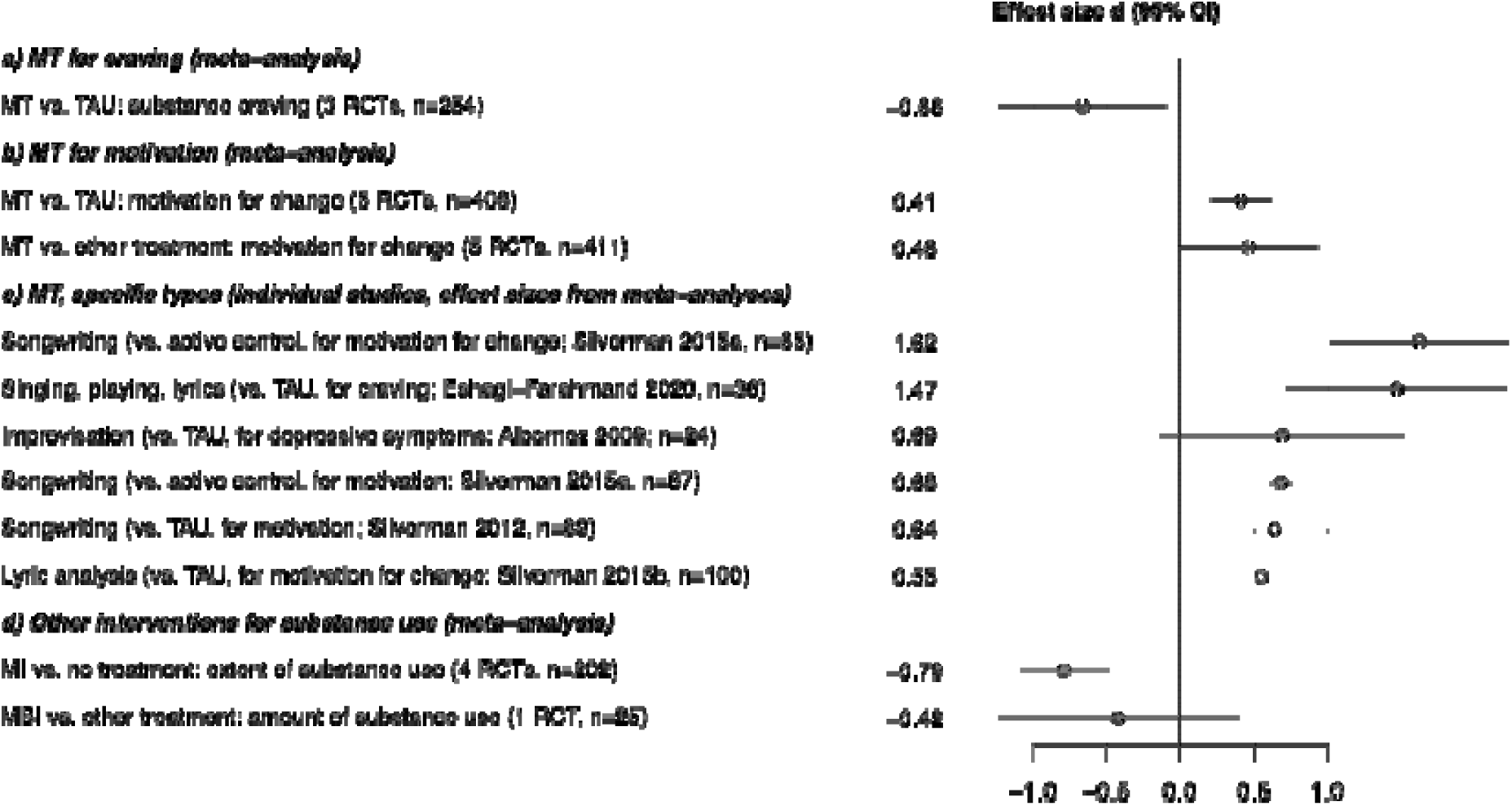
Effects of selected psychosocial interventions for substance use disorder. *Note.* Short-term effects of MT on craving, motivation, and mental health outcomes (a-c); effects of other interventions on substance use (d), from Cochrane reviews (all measured at end of intervention)^23,34,35^. Details on individual study labels (c) see Cochrane review^23^. MBI – mindfulness-based intervention; MI – motivational interviewing; MT – music therapy; RCT – randomised controlled trial; TAU – treatment as usual.

Various forms of MT have been found beneficial for people with substance use disorder*^23^*. Effects of MT may vary according to specific MT type (e.g. active, listening, combined; all provided in group settings; Fig. 1c). Additionally, effects may depend on participant characteristics such as SUD type, age, gender, and minority status. Participants in MT studies to date used most commonly alcohol (52%), followed by heroin (30%), prescription drugs (15%), cocaine/crack (2%), other substances (11%), or combinations of substances*^23^*. The FALCO trial will examine both active and listening MT groups and will include a broad spectrum of participants with various SUD.

Direct evidence on MT is almost exclusively focused on immediate and short-term effects, particularly on precursors of clinical outcomes such as motivation and craving. However, indirect evidence suggests that related interventions such as MBI*^34^* or MI*^35^* may have effects on substance use (Fig. 1d). There is therefore a need to examine long-term effects (1 year and beyond) on clinical outcomes, such as addiction severity and recovery. Yet the potential of MT as a part of *comprehensive rehabilitation* for people with SUD remains unexplored in a systematic way across multiple countries and treatment sites. No RCT to date has evaluated the long-term outcome of an active music group (AMG) or a music listening group (MLG) versus treatment as usual (TAU). Fig. 2 depicts the comprehensive approach of the FALCO trial: it will examine patient-relevant long-term clinical effects (right end of Fig. 2) as well as the shorter-term mechanisms and outcomes leading to them (middle sections of Fig. 2), considering the interplay of AMG and MLG with TAU (left end of Fig. 2).

**Figure 2.**
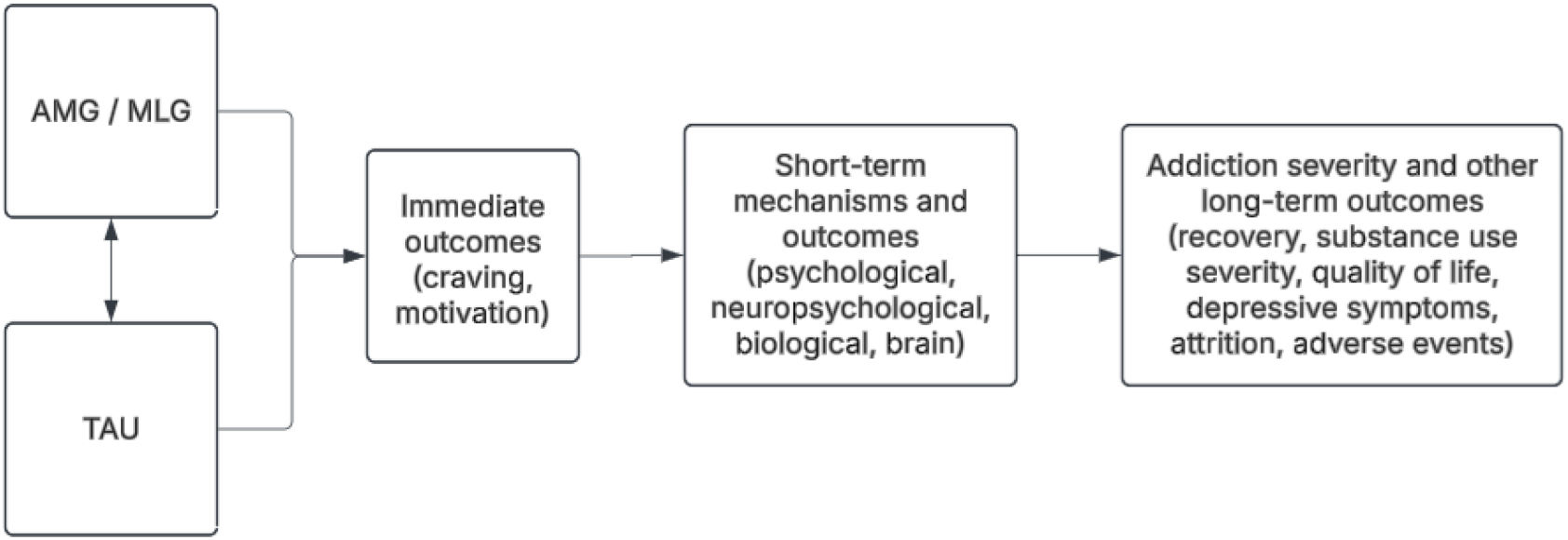
Logic model of interventions and outcomes to be tested in FALCO. *Note*. AMG – active music group; MLG – music listening group; TAU – treatment as usual.

## 2. Objectives and hypotheses

The primary confirmatory objective of the FALCO trial is to determine effects of AMG or MLG in addition to TAU versus TAU alone on addiction severity in people with SUD at one year after randomisation (Fig. 2, Fig. 3).

**Figure 3.**
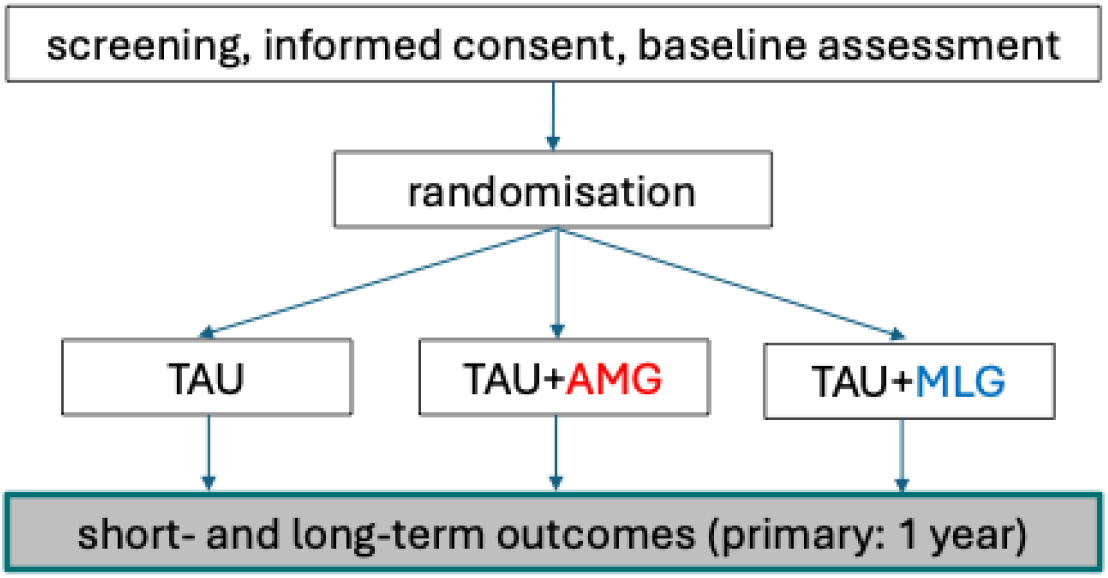
Overall design of the FALCO trial. *Note*. AMG – active music group; MLG – music listening group; TAU – treatment as usual

Secondary exploratory objectives are as follows:

- Determining the effects of AMG or MLG versus TAU on further long-term / downstream clinical outcomes, including recovery, quality of life, mental health outcomes, and adverse events. Evaluations will be performed at one year (primary endpoint) and two years.
- Determining the effects of AMG or MLG versus TAU on short-term / direct, mechanistic outcomes, which can be expected to change more quickly. These include psychological, cognitive, biological, and structural brain changes, which will be evaluated at three to 12 months.
- Determining the changes in immediate outcomes, motivation and craving, from before to after a session of AMG, MLG, or a psychosocial intervention session provided as part of TAU (e.g., mindfulness-based intervention, motivational interviewing, physical activity).
- Very long-term outcomes up to 10 years from randomisation. These will be collected from electronic health records without any active participation of the study participants.

We hypothesise that both AMG and MLG in addition to TAU will be superior to TAU alone with respect to long-term and short-term outcomes. Additionally, we hypothesise that immediate outcomes after individual sessions will differ between AMG, MLG, and TAU interventions, and that these will predict downstream short- and long-term outcomes.

An additional goal of the overall project is to examine behavioural and functional brain mechanisms affected by AMG and MLG versus TAU using specifically designed experimental paradigms combined with functional brain imaging. These studies, embedded within the FALCO trial, will be developed and reported separately, and are not detailed in this RCT protocol. Further studies embedded within the FALCO trial will concern process evaluations and user perspectives. Peer worker involvement enhances care in SUD treatment^18^, but its potential in MT is still unexplored. A broader goal of the FALCO project is to harmonise and standardise intervention protocols across Europe, while simultaneously preserving and enhancing tailoring to specific sub-populations, defined geographically, culturally, ethnically, demographically, or clinically, to ensure that interventions are widely accessible.

For clarity and conciseness, the present manuscript describes the methods for the primary and the main secondary outcomes. Embedded studies within a trial (SWATs) are described briefly in the main text but will be described more fully in separate protocols. These include hair biomarkers; blood biomarkers; brain imaging; immediate outcomes; process evaluations; machine learning analyses; and 10-year outcomes. Participation in hair and blood biomarkers, brain imaging, and process evaluations is optional for sites and participants.

## 3. Methods

### 3.1 Design

The FALCO trial is a parallel 3-arm assessor- and statistician-blinded pragmatic multinational RCT to evaluate the long-term effects of MT in individuals with substance use disorder. After the primary endpoint, an open-label extension will allow participants to choose their preferred intervention (Fig. 4). This will be important to sustain motivation to participate among those randomised to TAU. The outcomes at two years and beyond will therefore contain randomised (until 1 year) and non-randomised interventions (after 1 year). While this may reduce the observable effects, it is preferable to higher attrition, which would be a risk if not allowing this choice.

**Figure 4.**
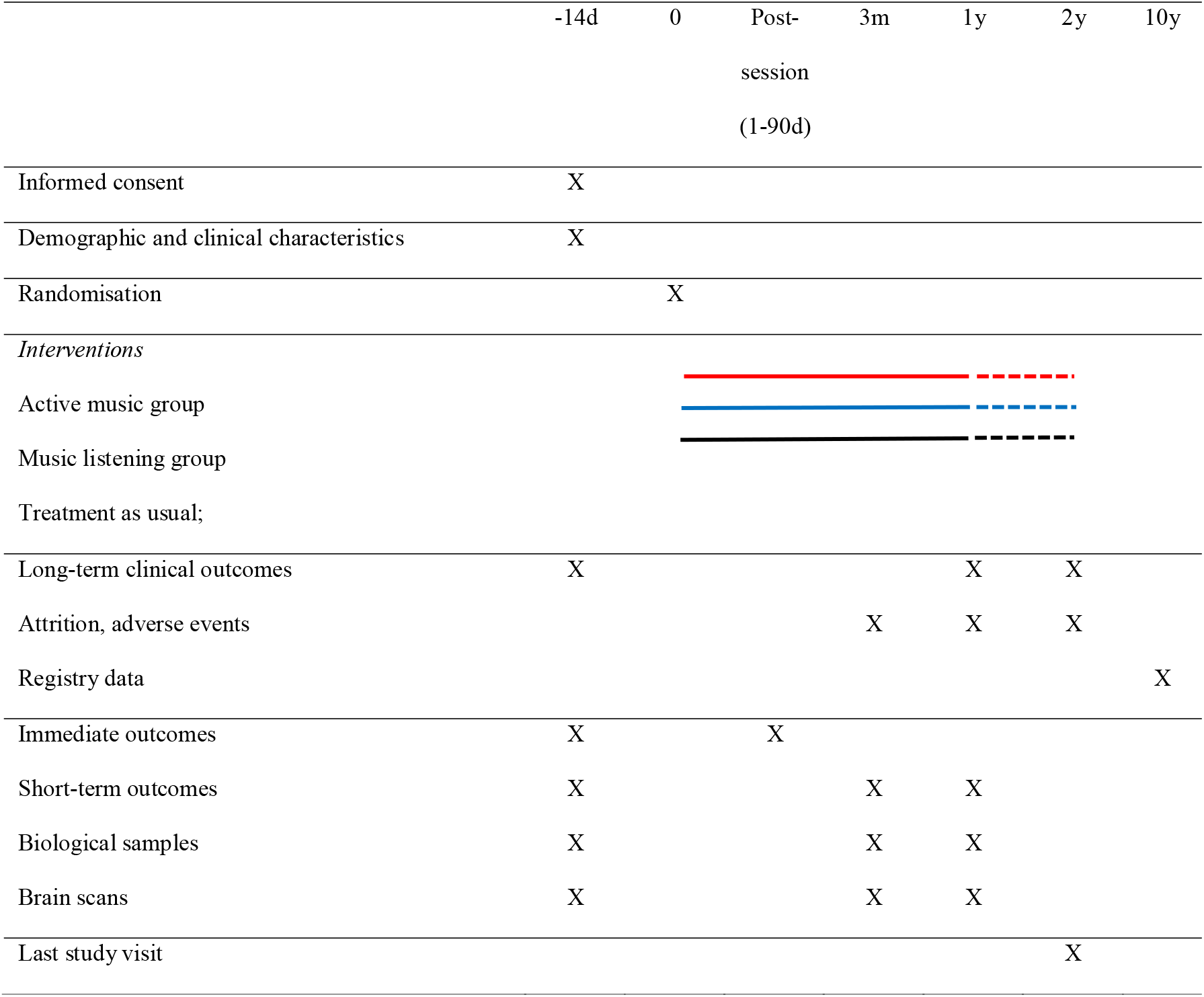
SPIRIT schedule of FALCO trial procedures. *Note.* d – day; m – month; y – year; solid line – participants receive randomised intervention; dashed line – participants choose preferred intervention regardless of study arm.

Participants will be randomised in a 1:1:1 ratio, stratified per site, with no further patient-related stratification levels, using a form of restricted randomisation (block randomisation with randomly varying block sizes; details documented separately to reduce predictability).

A concealed, computer-generated randomisation sequence was produced by a researcher with no direct contact with participants, and randomisation results will be administered centrally through a web-based system (REDCap; assignments cannot be accessed in advance) after informed consent and baseline assessment. Written informed consent will be collected by researchers, or clinical site staff who are trained in the study procedures; a model consent form is enclosed (Supplementary Material 1, SM1). This form has been translated into the relevant local languages of all participating countries by local research teams to ensure linguistic and cultural appropriateness.

The primary outcome and all other clinician-based measures will be conducted by blinded assessors. Blinding success will be verified by asking the assessor at the end of the primary endpoint assessment if they inadvertently found out the participant’s allocation, and if so, to state the condition they believe the participant was allocated to. Participants are instructed at the beginning of the interview to not reveal their allocation. Assessors are separate from therapists, work on separate units, and are not involved in any clinical team meetings. Statisticians conducting analyses will also be blinded, and report writing will be blinded until a draft is ready. For analysis, treatment arms will be coded with letters (A-C). An unblinded statistician who is not involved in the trial or the analyses will hold the code. Unblinding will occur after the finalisation of the statistical analysis plan and primary results.

Follow-up intervals follow consensus guidelines on core outcome sets for addiction*^36^*. Participation in the trial ends with the 2-year assessment (Fig. 4); a later follow-up will be based on registry data and does not require participation of former participants (Fig. 4). Protocol changes since the initial version are shown in Table 1, and an overview of study procedures with time required to complete can be seen in Table 2.

**Table 1.** History of protocol changes.

| Version number | Date | Type of change | Rationale |
|---|---|---|---|
| V1.15 (this version) | 28 October 2026 | Clarification of methods and procedures | Requests from journal |
| V1.12 | 19 February 2026 (posted on preprint server) | Correction of reference numbers and typos | -- |
| V1.10 | 19 January 2026 | Minor text revision | Requests from journal |
| V1.08 | 2 December 2025 (submission to journal) | Major text revision; revised inclusion criteria (clarifications; age limits changed to ≥18); revised assessment tools; no patient-related stratification variables in the randomisation | Homogeneity of sample; feasibility of recruitment and assessments |
| V0.2 | 12 March 2025 (revised Norwegian ethics submission) | Raclopride PET (Norway) removed; PET to be conducted only in Spain. Blood samples not taken in Norway, Israel, Italy | Not possible in these locations |
| V0.1 | 11 April 2024 (grant submission date) | -- | -- |

*Note.* All methodological changes listed in this table were approved by the relevant ethics committees and updated in the trial registry before implementation.

**Table 2.** FALCO assessment protocol with time needed to complete.

| Evaluation/test type | Name | Measure of / Type of measure or test | Items | Minutes @ baseline | Minutes @ follow up | Baseline | First session (pre/ post) | 3m | 1y | 2y |
|---|---|---|---|---|---|---|---|---|---|---|
| Baseline evaluation | **Demographics** | Interview | 10 | 10 | 0 | x | - | - | - | - |
|  | **Extended Barcelona Music Reward Questionnaire (eBMRQ)** | Musical hedonia | 24 | 7 | - | x | - | - | - | - |
|  | **Healthy - Unhealthy music scale** | Use of music | 13 | 4 | **-** | x | - | - | - | - |
|  | **Profile of Music Perception Skills (Mini-PROMS)** | Musical Skills | 4 | 15 | **-** | x |  |  |  |  |
|  | **WAIS Vocabulary** | IQ | 40 | 7 | 0 | x | - | - | - | - |
| Long-term outcomes | **European Addiction Severity Index (EuropASI)** | Addiction severity / Interview | 164 | 75 | 25 | x | - | - | x | x |
|  | **Substance Use Recovery Evaluator (SURE)** | Recovery / Self-report | 21 | 5 | 5 | x | - | - | x | x |
|  | **Drug Use Disorder Identification Test Consumption items (DUDIT-C)** | Drug use / Self-report | 4 | 2 | 2 | x | - | x | x | x |
|  | **Alcohol Use Disorder Identification Test Consumption items (AUDIT-C)** | Alcohol use / Self-report | 4 | 2 | 2 | x | - | x | x | x |
|  | **EQ-5D-5L** | Quality of life / Self-report | 5 | 5 | 5 | x | - | - | x | x |
|  | **Montgomery-Åsberg Depression Rating Scale (MADRS)** | Depressive symptoms / Interview | 10 | 5 | 5 | x | - | - | x | x |
|  | **Attrition** | Attendance data | 0 | 0 | 0 | x | - | x | x | x |
|  | **Adverse effects** | Interview | 0 | 0 | 5 | - | - | x | x | x |
| Pre-post session | **Importance, Confidence, Readiness Ruler (ICR)** | Motivation / Self-report visual analogue scale | 3 | - | 2 | - | x | - | - | - |
|  | **The Craving Scale** | Craving / Self-report visual analogue scale | 3 | - | 2 | - | x | - | - | - |
|  | **Inclusion of Other in the Self Scale** | Social connectedness / Self-report | 1 | - | 1 | - | x | - | - | - |
| Short -term | **The Craving Scale** | Craving / Self-report visual analogue scale | 3 | 1 | 1 | x | - | x | x | - |
|  | **University of Rhode Island Change Assessment (URICA)** | Motivation for change / Self-report | 32 | 10 | 10 | x | - | x | x | - |
|  | **Snaith-Hamilton Pleasure Scale (SHAPS)** | Anhedonia / Self-report | 14 | 5 | 5 | x | - | x | x | - |
|  | **Sleep Condition Indicator (SCI)** | Sleep quality / Self-report | 8 | 3 | 3 | x | - | x | x | - |
| Cognitive performance | **CANTAB - Rapid Visual Information Processing** | Sustained attention | - | 6 | 6 | x | - | x | x | - |
|  | **CANTAB - Spatial working memory** | Working memory | - | 10 | 10 | x | - | x | x | - |
|  | **CANTAB - Paired Associates learning** | Learning and memory | - | 12 | 12 | x | - | x | x | - |
|  | **CANTAB - Intra-Dimensional/extra-dimensional set-shifting** | Set-shifting | - | 12 | 12 | x | - | x | x | - |
|  | **Cognition of Social Interaction in Movies (COSIMO)** | Social cognition | - | 6 | 6 | x | - | x | x | - |
|  | **Test of Creative Imagery Abilities (TCIA), short version** | Creativity | - | 7 | 7 | x | - | x | x | - |
| Biological samples | **Hair samples** | Substance use | - | 5 | 5 | x | - | x | x | - |
|  | **Blood samples (5 sites: Vienna, Gdansk, Barcelona, Girona, Zurich)** | Brain pathology | - | 5 | 5 | x | - | x | x | - |
| Brain scans | **Magnetic Resonance Imaging (MRI, fMRI, DTI; 4 sites: Bergen, Vienna, Barcelona, Zurich)** | Brain structure, white matter connectivity, task-related functional activity | - | 60 | 60 | x | - | x | x | - |
|  | **Positron Emission Tomography (PET; 1 site: Barcelona)** | Brain dopaminergic metabolic activity | - | 100 | - | x | - | - | x | - |
| **Sum (not including blood samples and brain scans)** |  |  | **361** | **214** | **131** |  |  |  |  |  |

*Note*. Abbreviations see text.

### 3.2 Study settings and participants

We will enrol 600 patients with SUD in Europe and the Middle East (coordinated in Norway, initiated with Austria, Israel, Italy, Poland, Spain, Switzerland; Palestine added later; updated list see trial registration). Patients will be recruited directly through partner institutions that are connected to a SUD rehabilitation centre, as well as through advertisements in traditional and social media. User representatives will play an important role as advocates of the trial to spread the information in their formal and informal networks. In some countries, existing databases from general practitioners will also be used to aid recruitment. A site investigator enrols participants, confirms eligibility, obtains consent, oversees baseline assessments, and confirms readiness to be randomised. A site investigator who is not involved in follow-up assessments then performs randomisation and informs intervention providers.

#### Eligibility

Participants are patients seeking or receiving treatment at the recruitment site for an existing substance use disorder based on ICD-10 criteria of both sexes. Participants may begin as either in- or outpatients, continue as outpatients, and continue participation if re-admitted to inpatient care. Eligible participants must have any type of substance use disorder including polysubstance and alcohol use (ICD-10 F10-16 and F18), except nicotine use disorder (F17; may be present in addition to the above). Diagnoses are confirmed by the clinical research team at the site according to ICD-10 criteria. No minimum current severity is required. Because this study is focused on rehabilitation rather than detoxification, eligible participants will not currently be in a detoxification unit or programme, but in a rehabilitation setting. Participants may have comorbid mental disorders (e.g., anxiety, depression, trauma-related disorders), but no psychotic episode in the last three months. Eligible participants will be at least 18 years old on the day of randomisation, with no upper age limit. Many substance users now survive into old age due to improved care. Further exclusion criteria, as judged by the local clinical research team at the site based on the clinical interview, are an existing diagnosis of dementia, insufficient language skills to participate in treatment, hearing impairment that considerably impairs hearing of music played at a moderate volume (unless hearing is sufficiently compensated by a hearing aid), and is currently receiving MT or has received regular MT at planned and reoccurring intervals, during the past year.

In line with the intention-to-treat principle, participants who meet exclusion criteria after randomisation (e.g. those who relapse and need detoxification, develop a psychotic state, or need to pause interventions) will remain in the study.

#### Baseline evaluation

Before randomisation, participants will complete demographics as described above; written informed consent; baseline assessments of outcomes described below; and the following assessments (Table 2): To assess music-related characteristics of participants, we will use the extended version of the Barcelona Music Reward Questionnaire (eBMRQ) measuring musical hedonia; the Healthy-Unhealthy Music Scale (HUMS) measuring risk-related or protective music engagement^10^; and a short version of the Profile of Music Perception Skills (Mini-PROMS) measuring musical skills^39^. A premorbid verbal intelligence quotient (IQ) estimate will be assessed using the vocabulary test of the Wechsler Adult Intelligence Scale 4^th^ version (WAIS-IV). This test includes 30 words to be defined (depending on the language) and takes approximately 5-10 minutes to complete (Table 2)^40^.

### 3.3 Interventions

The interventions of interest in the FALCO trial (AMG, MLG) are non-pharmacological psychosocial interventions. TAU will serve as the comparator, allowing the new interventions to be evaluated against routine, everyday practice.

Participants will continue to receive TAU as offered locally, which may include non-pharmacological and pharmacological interventions. These interventions are not experimental and may differ between sites and participants, but will be balanced between study arms through randomisation. There are no minimum expected compounds of TAU. TAU interventions received by participants and any changes in these interventions will be registered at each assessment time point (0, 3, 12 months) using a specifically devised form (SM2). Participants are asked not to participate in MT outside the study for 12 months. Self-directed music activities undertaken by participants outside of intervention sessions are permitted and will be recorded; in a brief questionnaire, participants will be asked to specify the type (singing or playing an instrument; on their own vs. with others), duration, and frequency of active music-making over the course of the last three months. Any changes in such activities will also be recorded at the same time points.

Both AMG and MLG will be conducted by appropriately qualified music therapists. Eligible therapists are graduated from an academic MT training program (at master’s level or equivalent) and have at least one year of full-time equivalent experience working as a music therapist in a group setting with persons with SUD. Professional registration is required if applicable in the given country (e.g. Austria). AMG and MLG will be provided in groups, with weekly sessions lasting up to 90 minutes. An additional period of 15 minutes will be provided pre-group for arrival of and informal exchange between participants and for the therapist to prepare the session, and 15 minutes post-group for the therapist to clear up the room and for further informal exchange between and departure of the participants. The length of sessions will be determined by the interventionists, based on patients’ needs, clinical judgement, local settings etc. Group sizes will be around 6 participants in both intervention arms. Group sessions may be supplemented with individual sessions if needed (e.g., to prepare for groups). MLG requires a stereo system and music recordings (e.g. via a streaming platform). AMG requires a variety of musical instruments*^41^*. A list of included instruments is provided in Table 3.

**Table 3.** List of instruments required for music groups.

| **Category** | **Type of instrument** | **Number of instruments** |
|---|---|---|
| **Access to streaming platform for music listening** |  | 1 |
| **Keyboards** | Electric/acoustic piano | 1 |
| **Strings** | Guitar (steel/nylon stringed) | 2 |
|  | Harp | 1 |
|  | Ukulele | 2 |
| **Drums** | Hand drum | 3 |
|  | Djembe | 3 |
|  | Bongo | 2 |
|  | Tambourine | 2 |
|  | Cajon | 1 |
|  | Darbuka | 2 |
| **Auxiliary Percussion** | Wood block | 2 |
|  | Hand jingle bells | 2 |
|  | Guiro | 2 |
|  | Wood claves | 3 |
|  | Shaker | 4 |
| **Melodic Percussion** | Wood xylophone | 2 |
|  | Metallophone | 1 |
|  | Slit drum | 1 |
| **Wind instruments** (if disinfection is feasible) | Flute | 2 |
|  | Harmonica | 2 |
|  | Whistle | 2 |
|  | Melodica | 2 |
| **Band-oriented instruments** | Electric bass guitar | 1 |
|  | Synthesizer | 1 |
|  | Drum-set (or electric drum-set) | 1 |
|  | Electric guitar | 1 |
| **Other** | Microphone | 2 |
|  | PA system with mixer | 1 |
|  | Amp | 2 |
|  | Computer/laptop<br>(with software licenses as needed) | 1 |
|  | Mobile recording device<br>(and audio interface device as needed) | 1 |

*Note.* The type and the number of instruments in the table are suggestions based on consortium consensus. It is advisable to check with therapists at each site which specific items of equipment are needed so that the music interventions can be delivered in accordance with therapists’ specific skills and cultural contexts (e.g., advanced electronic equipment and software will only be needed if therapists are accustomed to using it in their work).

Both AMG and MLG will be provided for 12 months as randomised. From one-year post-randomisation, participants will be allowed to choose their preferred intervention regardless of study arm (Fig. 4). Giving participants the opportunity to attend music groups after one year serves important ethical and practical purposes (including encouraging continued involvement and reducing attrition), while also retaining the integrity of the original randomised comparison until the primary endpoint.

AMG and MLG will be provided following intervention guidelines developed in the initial phase of this study (SM3). These specify intervention elements while maintaining flexibility for cultural adaptations, local circumstances and participant needs. Interventionists will receive remote training in application of these guidelines, and continuous peer supervision through fortnightly online meetings between researchers and interventionists across sites to ensure fidelity and intervention quality. Study-specific MT training comprises a 3-hour online session for music therapists before starting to deliver sessions. Fortnightly online peer supervision meetings are held for one hour each, with a shared minutes document ensuring that relevant topics are accessible to all interventionists throughout the study period. During training and supervision, researchers responsible for treatment quality and fidelity routinely check entries in online session forms, session videos, and interventionist contributions in supervision meetings to make sure that sessions are implemented adequately. Where available, peer workers (individuals with lived experience of a SUD who have undergone specific training to offer support to people with SUD alongside professionals; see section 6) are recruited locally to act as co-providers of interventions. They also undergo study-specific training before co-delivering sessions and are also required to regularly attend peer supervision meetings.

Registers will be used to routinely record provision of group MT sessions (AMG, MLG) and participants’ attendance in those sessions (regardless of current in- or outpatient status). Fidelity will be assessed by analysing self-report forms completed by interventionists after each session and evaluating video recordings of sessions. Details of treatment fidelity, such as the sampling strategy and proportion, selection and blinding of raters, assessment of inter-rater reliability, and specific measures of engagement will be explained in a separate manuscript.

### 3.4 Primary outcome and other long-term clinical outcomes

For all outcomes, availability of validated translations was a guiding principle. We used established core outcome sets to select measures and time points. In cases where a validated translation was not available, we used standard methods (translation and independent back translation, including cultural adaptation of item wording as applicable). A dedicated working group provides assessor training, verifies reliability, and supervises assessments throughout the study. All assessors underwent internal online assessment training and certification (online meetings with A.H.E. and J.B.). To keep the assessment burden manageable and to avoid participant fatigue or distress, sites are encouraged to adjust baseline measurement according to their possibilities (e.g., provide breaks, water, split assessment into two meetings). For details on monitoring of these outcomes as well as follow-up and retention strategies, see SM4.

This section includes key outcomes that are clinically relevant for patients and that tend to take an extended time to emerge. All long-term outcomes described in this section are survey-based (clinician rating or self-report) and assessed at baseline, at one year, and at two years unless stated otherwise.

*The primary outcome* is <u>addiction severity</u>, which will be assessed using the European Addiction Severity Index (EuropASI). The survey covers a broad concept of addiction, including changes that typically require a long time, across seven domains (general medical condition, professional and financial situation, substance use, other drug consumption, legal problems, family and social relations, psychological condition), and has been adapted specifically for the use in European contexts*^42^*. This scale is clinician-based and widely used and recommended*^6^*, involving a 20-25 minute interview (45-75 minutes at baseline). The scale includes 164 items*^42^*, with a fine-grained Composite Score of 0 (best) to 1 (worst) and is freely available in many languages*^6^*. It has demonstrated good psychometric properties across languages. The primary variable is the score of the EuropASI representing each participant’s substance use severity: the alcohol composite score, drug composite score, or their average, for those with alcohol use disorder alone (ICD-10 F10), drug use disorder alone (F11-F16 or F18), or a combination (F10 and any of F11-16 or F18). It will be measured at 12 months, with a tolerance window of ±1 month. The two primary estimands are the treatment effect of AMG plus TAU versus TAU alone and MLG plus TAU versus TAU alone, in adults with SUD, on addiction severity (EuropASI alcohol/drug composite score endpoint, adjusted for baseline) at 12 months from randomisation, in the intention-to-treat population as randomised, regardless of intervention discontinuation, relapse, hospitalisation, incarceration, receipt of non-study MT or treatment crossover. This outcome does not exist in the event of death. Handling of missing data is described in section 3.9.

*Key secondary outcomes* are the remaining composite scores and time points of the EuropASI and other long-term clinical outcomes relevant to the studied group. For outcomes measured at two years, it is important to consider that these may be affected by preference-based interventions; they cannot be interpreted as preserving the original randomised comparison once participants choose their intervention. The estimands for these outcomes can be described as the effect of referring adults with SUD to AMG or MLG a year earlier than in TAU alone, where access to AMG or MLG is delayed.

<u>Recovery</u> will be assessed using the Substance Use Recovery Evaluator (SURE). Similarly to the EuropASI, the scale measures a construct that is broader than the mere amount or extent of substance use. It captures five factors (substance use, material resources, outlook on life, self-care, relationships) using 21 items. The possible range is 21 to 63. It is patient-reported and was developed with strong user involvement*^43^*. This scale was developed in collaboration with users and practitioners*^43^*, is recommended in SUD core outcome sets*^36^*, and freely available in many languages. However, as it has not been used in many intervention studies yet, its sensitivity to change is less well established than that of the EuropASI, and it was therefore not selected as primary.

<u>Substance use severity</u> will be assessed using the consumption items from Drug/Alcohol Use Disorders Identification Tests (DUDIT-C/AUDIT-C). DUDIT-C/AUDIT-C are also assessed at three months, as the scales are sensitive to short-term changes as well. These scales are patient-reported short screening tools that can be used repeatedly and have been used in long-term follow-up studies of SUD*^44^*. The possible range is 0 to 12 for AUDIT-C, while DUDIT-C range from 0 to 16. They are recommended in SUD guidelines*^6^* and rate for DUDIT-C the use as 0 for self-reported abstinence to ≥ 7 for heavy use*^44^*, and for AUDIT-C 0 for no alcohol use and ≥ 3/4 (women/men) as alcohol misuse*^45^*.

<u>Quality of life</u> and <u>depressive symptoms</u> will be assessed using the EuroQol (EQ-5D)*^46^* and the Montgomery-Åsberg Depression Rating Scale (MADRS)*^47^*, respectively. The EQ-5D is a patient-reported scale with 5 items plus a visual analogue scale. It is widely used across all clinical domains, ranges from “less than 0” (worse than death) to 1 (best), and is freely available in many languages with country-specific value sets. The possible range is 5 to 25 on the items, while the visual analogue scale ranges from 0 to 100. It is commonly used as a basis for economic evaluations (https://euroqol.org). Depression is a common comorbidity that has successfully been addressed by MT in SUD*^23^*. The MADRS is a clinician-administered ten-item interview measuring symptoms of depression. The scale is well suited to assess severity of depressive symptoms (rating symptoms on a 7-point scale), and is a preferred assessment tool for clinical trials, due to its superior sensitivity to change. The possible range is 0 to 60, where higher scores indicate increasingly severe depressive symptoms.

<u>Attrition and adverse effects</u> will be assessed at three months, one year and two years. Regarding attrition, clinicians will judge whether a participant is still participating, has decided to withdraw from the study, or contact has been lost. Importantly, participants will not be regarded as withdrawn if they decide to withdraw from interventions or from some assessment procedures. Attempts will be made to follow up all participants unless they state that they would like to withdraw from all study procedures or repeated attempts to contact them are futile (see section on retention below).

In line with ICH E2A guidance*^1^*, we define adverse events (AEs) as any untoward medical occurrence in a participant, related or unrelated to interventions, and serious adverse events (SAEs) as any AE that requires inpatient hospitalisation or prolongation of existing hospitalisation, results in persistent or significant disability/incapacity, or is a congenital anomaly/birth defect (in a participant’s offspring). Since little is known about any SAEs resulting from music interventions, site investigators will first ask participants in all study arms at each visit to describe any adverse events, and will then rate whether these were serious or non-serious and related or non-related according to the definitions above. This will then be adjudicated centrally by the research team. SAEs will be reported immediately. Re-hospitalisations, relapses, and worsening of mental health conditions are expected due to the condition of the participants and will be monitored by site investigators, followed up by local clinical teams, and reported. Potential harms from music interventions may include triggering craving, distressing memories, interpersonal conflict, overstimulation, hearing discomfort, and reinforcement of unhealthy music-use patterns. These will usually be transient and form a part of the therapy process. Therapists are trained to work with such events and will discuss them in supervision and in their clinical team (see section 3.3 Interventions). Assessors will also ask about such AEs during each study visit.

Patients’ participation in the study ends after two years. Outcomes up to 10 years from randomisation will be collected from electronic health records and similar public records (SM10).

### 3.5 Exploratory short-term outcomes (3 and 12 months)

Short-term outcomes are secondary outcomes and will be assessed at baseline, at three months and at one year (Table 2).

#### 3.5.1 Questionnaires

<u>Motivation for change</u> will be assessed with the University of Rhode Island Change Assessment (URICA)*^48^*. This patient-reported scale is more comprehensive than the very brief ICR that is used as immediate outcome. URICA measures the extent to which individuals identify with different stages of change (precontemplation, contemplation, action, maintenance) across 32 items. Each subscale consists of eight items using 5-point Likert scales ranging from 1 to 5, resulting in scores ranging from 8 to 40. Subscale scores can be used to trace changes in attitudes related to the specific stages of change. The subscales are combined arithmetically (Contemplation + Action + Maintenance - Precontemplation) to yield a Readiness to Change score, with a possible range of -16 to 112 and higher scores indicating greater readiness to change. The scale has been used in previous MT trials*^23^*.

<u>Craving</u> will be assessed using The Craving Scale*^49^*. This is a patient-reported scale including three items that has been used in previous SUD-studies *^50–52^*. Scores range from 0-9, with high scores indicating higher cravings.

<u>Anhedonia</u> will be assessed with the Snaith-Hamilton Pleasure Scale (SHAPS)*^53^*, a widely used patient-reported scale that measures the capacity to derive pleasure from four different hedonic experiences: interest/pastimes, social interaction, sensory experience, and food/drink. The scale has 14 items, each scoring from 0 to 3, producing a sum score ranging from 0 to 42*^54^*. Higher scores represent higher anhedonia level and a lower hedonic tone. The scale has been widely used with populations presenting different mental health conditions, including SUD patients*^55^*.

<u>Sleep quality</u> will be assessed with the Sleep Condition Indicator (SCI)*^56^*. The SCI includes eight questions covering various sleep aspects, such as sleep onset delay, night-time awakenings, extent of the problem and its effect on daytime activities, mood and relations. Responses are scored from 0 (poor state) to 4 (no/minor problems). The possible range is 0 to 32., with a total score ≤16 indicating probable insomnia. Maximum score is 32. By using the SCI, we aim to identify sleep issues and understand participants’ sleep quality, contributing to sleep research.

#### 3.5.2 Biological samples

##### Hair samples

Objective measures of substance use from hair testing have been shown to reliably determine substance use at a higher sensitivity compared to self-reports and are recommended to complement substance use interviews and questionnaires in clinical studies*^57^*. These measures are suitable to detect functionally relevant changes in individual substance use across time, as it was shown for cocaine use*^58^*. Hair biomarkers will be assayed for ≈100 substances, including illegal substances, alcohol use intensity, and psychotropic medications (details see SM5; further details will be described in separate protocols).

##### Peripheral (blood-based) markers of active brain pathology

The recent introduction of new-generation immunoassay methods allows the reliable quantification of structural brain markers in peripheral matrices such as blood. Neurofilament light chain (NfL), a neuron-specific cytoskeletal component, and glial fibrillary acidic protein (GFAP), a protein involved in maintaining astrocyte integrity, are both released in extracellular matrices after endogenous (e.g. neuro-inflammation and -degeneration) or exogenous (e.g., traumatic brain injury, intoxication) neuronal impairment*^59^*. Specifically, given its sensitivity to a wide range of neuropathological alterations, NfL has been suggested for the use in psychiatric practice – specifically in addiction medicine – as a highly sensitive, even though unspecific, tool to quantify active brain pathology*^59^* (details see SM6; further details will be described in separate protocols).

#### 3.5.3 Neuropsychological tests

##### Cognitive function

Cognitive function is often affected through substance use, but impairments are partially reversible in SUD*^58,60,61^*. Baseline verbal IQ assessment using the WAIS Vocabulary subtest (see above) will be conducted before these tests and will guide their interpretation. Cognitive performance will be assessed on tablets or on desktop computers with four tests from the Cambridge Neuropsychological Test Automated Battery (CANTAB)*^62^*. CANTAB tasks are well-grounded in neurobiology-informed cognitive concepts; they have cross-species translational validity and are sensitive to psychopharmacological effects*^63^*. All of the selected CANTAB tasks have been proven to be highly sensitive to cognitive impairments in different SUD populations*^64–71^*.

The dependent variables of the CANTAB tasks will be:

1. <u>Rapid Visual Information Processing (RVP):</u> This task focuses on sustained attention by assessing the discrimination performance A’ (a signal detection measure of target sensitivity integrating hits and false alarms) and total hits to assess attentional capacity. Median response latency (another psychomotor speed index), response bias B” (also known as acquiescence bias), and total false alarms will be used as measures of impulse control*^72^*.
2. <u>Spatial Working Memory (SWM):</u> This task mainly assesses visuospatial working memory. Number of total errors test the capability to retain spatial information and to manipulate remembered items in working memory. Additionally, applied heuristic strategies in this task are assessed by a strategy score reflecting the updating component of executive functions*^73^*.
3. <u>Paired Associates Learning (PAL):</u> This task measures visuospatial declarative learning and memory. The main outcomes are adjusted for total of errors and adjusted total of trials. The number of correctly located patterns after the first presentation is additionally assessed as a visuospatial working memory component*^74^*.
4. <u>Intra-Dimensional/Extra-Dimensional Set-Shifting (ID/ED):</u> This test covers rule acquisition and reversal, featuring visual discrimination, and attentional set-shifting, analogous to the Wisconsin Card Sorting Task*^75^*. Performance will be assessed by the number of trials (adjusted to the number of completed stages), the total number of errors (adjusted to the number of completed stages), the errors made up to the extra-dimensional shift (Pre-ED errors) and the errors made at the extra-dimensional stage of the task (ED errors)*^70^*.

##### Social cognition

The video-based online test COgnition of Social Interaction in MOvies (COSIMO; https://cosimo-project.com/) will be employed to measure <u>cognitive empathy and mental perspective-taking</u>. COSIMO has high ecological validity, is short, highly feasible (as it can be performed in a standard browser), and taps into potential core mechanisms of MT such as social interaction and social reward. Importantly, this test is well correlated with the Movie for the Assessment of Social Cognition (MASC) (r = .66, p < .001, n=130), which has been shown to be sensitive in SUD populations*^76^*, but being much shorter (5 min instead of 25 min). Performance will be assessed as number of correct answers.

##### Creativity

A shortened version of the Test of Creative Imagery Abilities (TCIA)*^77^* will be used to measure <u>creative imagery abilities</u>. Participants will be asked to view one by one simple graphic images, generate transformation ideas, and draw their favourite idea on paper with no time limit, with the possibility of elaborating or changing the selected image as they wish to create something more original. After each drawing, they will be asked to write what their drawing represents. The level of creative imagery ability will be measured by scoring the drawings according to three characteristics: vividness, originality, and transformative ability. We will administer a different set of items with a similar level of difficulty at each time point to prevent learning effects.

#### 3.5.4 Structural brain imaging

Structural brain imaging will include morphometric analyses from MRI, cortical folding and thickness, and structural connectivity from MRI-DTI (details see SM7; further details will be described in separate protocols). Functional brain imaging sub-studies using functional MRI and positron emission tomography (Table 2) in conjunction with specifically designed cognitive tasks will be described separately (see below).

### 3.6 Immediate outcomes

Exploratory immediate (post-session) outcomes include craving, motivation, and social connectedness based on brief self-report questionnaires. Comparing AMG and MLG to psychosocial interventions provided as part of TAU, this SWAT aims to bridge between results of previous RCTs showing immediate effects*^23^* to the longer-term outcomes addressed in this trial (SM8; further details will be described in separate protocols).

### 3.7 Participant timeline

The SPIRIT*^78^* schedule for the FALCO trial is shown in Fig. 4. Patients will be randomised following a baseline assessment and then followed up after initial intervention sessions, at three months and at one year. This concludes the main period of participation in the study. A long-term open-label extension will follow, in which participants will be provided with the opportunity to choose among available interventions (dashed lines in Fig. 4), with a last long-term follow-up visit at two years post-randomisation. This will conclude the patients’ participation.

### 3.8 Sample size

The trial is powered with 80% to answer the two main hypotheses of whether AMG and MLG are better than TAU alone regarding addiction severity at 12 months. The minimal clinically important difference for the alcohol/drug composite of the EuropASI is unknown, but from similar studies, an effect size of d≥0.4 is realistic: effect sizes in this range have been found in RCTs of MT and other psychosocial interventions for SUD*^23,34,35,79^*. This corresponds to a raw mean difference of 0.1 with a SD of 0.25, which is typically seen in studies using this score*^80–82^*. Moderate clustering by therapy group (ICC=0.05; group size 6) leads to a design effect of 1.25 (increasing the required sample size). With Bonferroni-adjusted significance level for two tests, this yields a required total sample size of 450 participants with valid data on the primary outcome (an independent samples t-test with two-sided significance level of 2.5% requires 120 subjects per group to achieve 80% for d=0.4; multiplying this number with the design effect stated above this leads to 150 subjects per group or 450 across all 3 arms). To compensate for ≤25% attrition, we will aim to recruit and randomise 600 participants (Fig. 5). The power calculation may be conservative because not all arms have data nested by therapy group and because baseline adjustments were not included in the power calculation.

**Figure 5.**
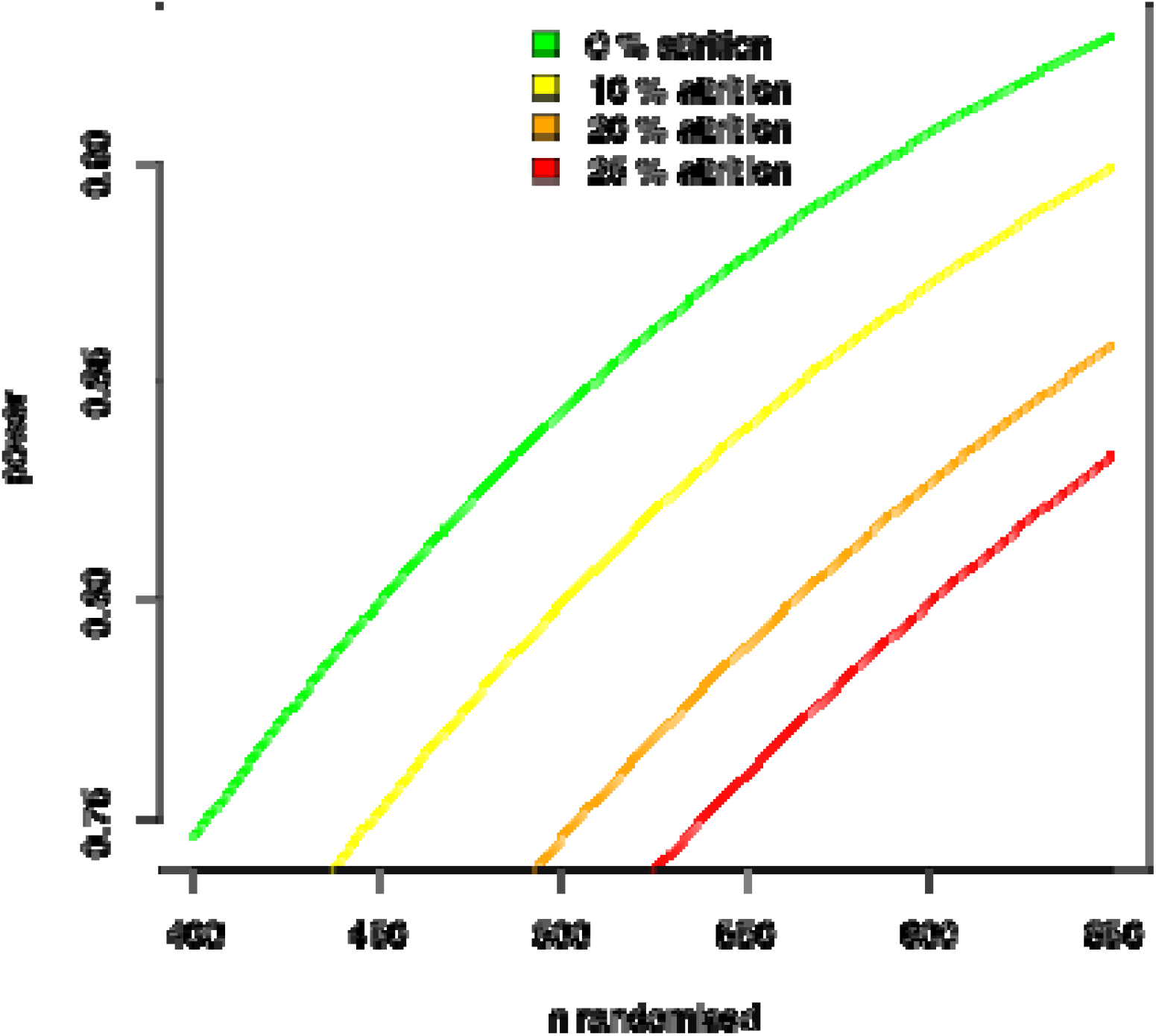
Test power of the FALCO trial for the primary endpoint under different attrition scenarios.

### 3.9 Statistical analyses

Intention-to-treat (analysing all participants as randomised) will be the guiding principle for the primary analysis; per-protocol analyses (interventions as actually received) will be added as a sensitivity analysis; see definition of analysis populations below. For the primary endpoint and other continuous outcomes measured at more than one time point (including baseline), we will use linear mixed-effects models (e.g., ANCOVA with the treatment contrasts AMG vs TAU and MLG vs TAU as fixed effects; country/site as nested random effects; and adjustment for baseline). The general significance level will be set to 5% (two-sided); the primary outcome will be Bonferroni adjusted to ensure a two-sided 5% level across both main comparisons (AMG vs. TAU; MLG vs. TAU). The effect estimates for the primary endpoint will be the difference in mean EuropASI drug/alcohol composite at 12 months between AMG and TAU, and analogously between MLG and TAU, with two-sided 97.5% confidence intervals to reflect the Bonferroni adjustment. Secondary outcomes will be exploratory.

Analysis populations are defined as follows:

– Intention-to-treat – including all participants as randomised, regardless of whether they were later found ineligible, did not adhere to the protocol or never attended an intervention session.
– Per-protocol – including those who attended at least 20 sessions over 12 months (i.e. 50% of an assumed realistic maximum of 40 sessions), with no additional criterion for treatment fidelity.
– Safety – those who attended at least one intervention session over 12 months.

We assume missing data to occur either missing completely at random (e.g. death) or missing at random (e.g. withdrawal/loss of contact). Strategies to prevent this second type of missingness are described above and in SM4. It may occur more frequently in TAU due to less frequent contact and loss of motivation. The mixed-effects models we will use are robust to data missing at random. To verify this assumption, we will compare baseline characteristics of those with missing with those of complete primary outcome. We do not plan multiple imputation.

Prespecified subgroup analyses will include SUD type (drug, alcohol, both); gender; and initial treatment setting (in- or outpatient).

To avoid conflicts of interest and to ensure the reproducibility of the statistical analyses, a comprehensive SAP will be developed. The SAP will contain detailed information about the principles, the assumptions and the description of the planned analyses as well as the way of result presentation using dummy tables and figures. The description of planned analyses will contain end points of primary and secondary analyses, methods of primary and secondary analyses, sensitivity analyses, significance levels and the handling of missing data. The SAP is accepted and signed by the Principal Investigator (PI), the trial statistician, and the quality control statistician.

#### 3.9.1 Analyses using machine learning (ML)

This combination of supervised and unsupervised ML approaches (SM9) complements the statistical trial analyses by focusing on prediction and, thus, on out-of-sample inference (generalisation to unknown data) using flexible, complex, high-dimensional, multivariable, yet robust, and credible models rather than on in-sample inference*^83,84^*. Therefore, this analysis will not be constrained or biased by assumptions about variable interactions, variable scales, or model oversimplification^58,59^.

## 4. Data collection and management

### Recruitment and retention strategies

Recruitment of participants will run in parallel at all sites until recruitment ends. An adequate time window of two years has been allotted to ensure the recruitment target can be reached in a collaborative effort between the sites. Re-allocation of resources from under-recruiting to over-recruiting sites is possible if needed. Retention is crucial in this study because attrition (from TAU) is known to be high in SUD (depending on substance type, e.g. stimulants vs. opioids vs. alcohol; comorbid mental disorders; and social status)*^3,85^*. This is also a particular challenge in studies of inaccessible clinical populations, such as individuals with SUD*^86^*, potentially leading to practical, ethical, and financial consequences. The extensive amount of time and resources needed for retaining participants in longitudinal studies should not be underestimated. A combination of strategies concerning study design and participant requirements, staff requirements, and technical strategies are of similar value, and their advantages will be most effective when used combined*^87,88^*. Staff will need both systematic and relational skills to uphold contact over time. We will use evidence-based strategies that can help reduce attrition, such as involving family members where possible*^85^*. At mid-recruitment, an analysis of retention rates will inform the final number of participants to be recruited to reach the target valid sample (see 3.8 Sample size above).

### Data management

We will use the widely used REDCap system, installed on NORCE servers (https://redcap.norceresearch.no/redcap/). The server uses the highest security standards including multi-factor authentication. Main outcomes will be captured using Electronic Case Report Forms (eCRFs) directly in REDCap. Some additional outcomes will be captured using paper CRFs (immediate outcomes, treatment fidelity) or other systems (computerised neuropsychological tests). User rights administration will use predefined roles (site manager, data entry person) and will be handled centrally. Data entry to REDCap is possible from any device with an internet connection. A central system for storage and analysis of other data (brain scans, intervention videos) is hosted by UIB (https://www.uib.no/safe), also with multi-factor authentication and user rights management as described above. Data export and preparation will be scripted in R for reproducibility and conducted centrally by NORCE. Data cleaning will be done by NORCE in collaboration with clinical sites at regular intervals. Videos, linkage keys, and pseudonymized data will only be available on SAFE, as described in the Data Management Plan which has been agreed by all partners. Personal identifiers and registry linkage keys will be maintained in separate access-restricted files. Intervention video recordings will be used solely for treatment fidelity monitoring, stored in encrypted repositories and accessible only to authorized personnel. For cross-national data management, all sites will comply with applicable national regulations and European General Data Protection Regulation (GDPR) requirements. Sharing of anonymised data is described in section 8.

## 5. Monitoring, harms and auditing

The monitoring bodies in FALCO are divided into several monitoring and advisory boards for quality assurance and appropriate risk management (Table 4).

- *Data Monitoring and Ethics Committee (DMEC):* The DMEC consists of a small number of members, at least three people with strong methodological and clinical expertise (at least 2 clinicians with expertise in SUD interventions and assessments; 1 statistician with expertise in non-pharmacological RCTs), who are independent of the trial (i.e. not involved with the trial in any other way; no competing interest that could impact on the trial; not affiliated with any of the institutions involved in conducting the trial). Any competing interests, both real and potential, will be declared/disclosed. The DMEC charter has been distributed to all DMEC members and is available on request from the corresponding author. The quorum for decisions is >50%; the DMEC chair will check with any absent members after meetings where recommending major action was considered. The DMEC recommends to the SSC via the trial office (Table 4). The DMEC had the initial protocol review meeting in 2025 and receives regular unblinded summaries of recruitment, attrition, uptake of interventions, unforeseen events, adverse events, and immediate information on any serious adverse events from the DMEC statistician. Meetings with the DMEC are held twice a year and consist of an open and a closed part. If any ethical or safety issues arise, the DMEC will recommend appropriate countermeasures to the study team.
- *Expert Advisory Group (EAG):* The EAG will consist of a European HTA body (Austrian Institute for Health Technology Assessment, partner in FALCO), 1-2 policy experts, 2-4 user representatives, and 5-6 experienced practitioners and researchers in the SUD field. It will act as an advisory body for the overall study implementation and will guide all user involvement strategies and activities within the study; meetings (in-person and online/hybrid) will take place 4-6 times each year.
- *Local advisory groups (LAGs):* 1 coordinator plus 4-8 user representatives, stakeholders and professionals from the field; will respond to the respective local circumstances of the RCT with regard to treatment practices, composition of the patients regarding (cultural) background, language, etc.
- *Study Steering Committee (SSC):* chaired by NORCE and comprised of Work Package (WP) leads and co-leads and one elected member of the Early Career Researchers (ECRs); supervisory body ensuring successful execution of the FALCO project, including suggesting the addition or removal of sites.
- *General assembly (GA):* chaired by NORCE, will consist of representatives from the whole consortium, with one representative from each partner having the right to vote; ultimate decision-making body about the FALCO project implementation.

**Table 4.**
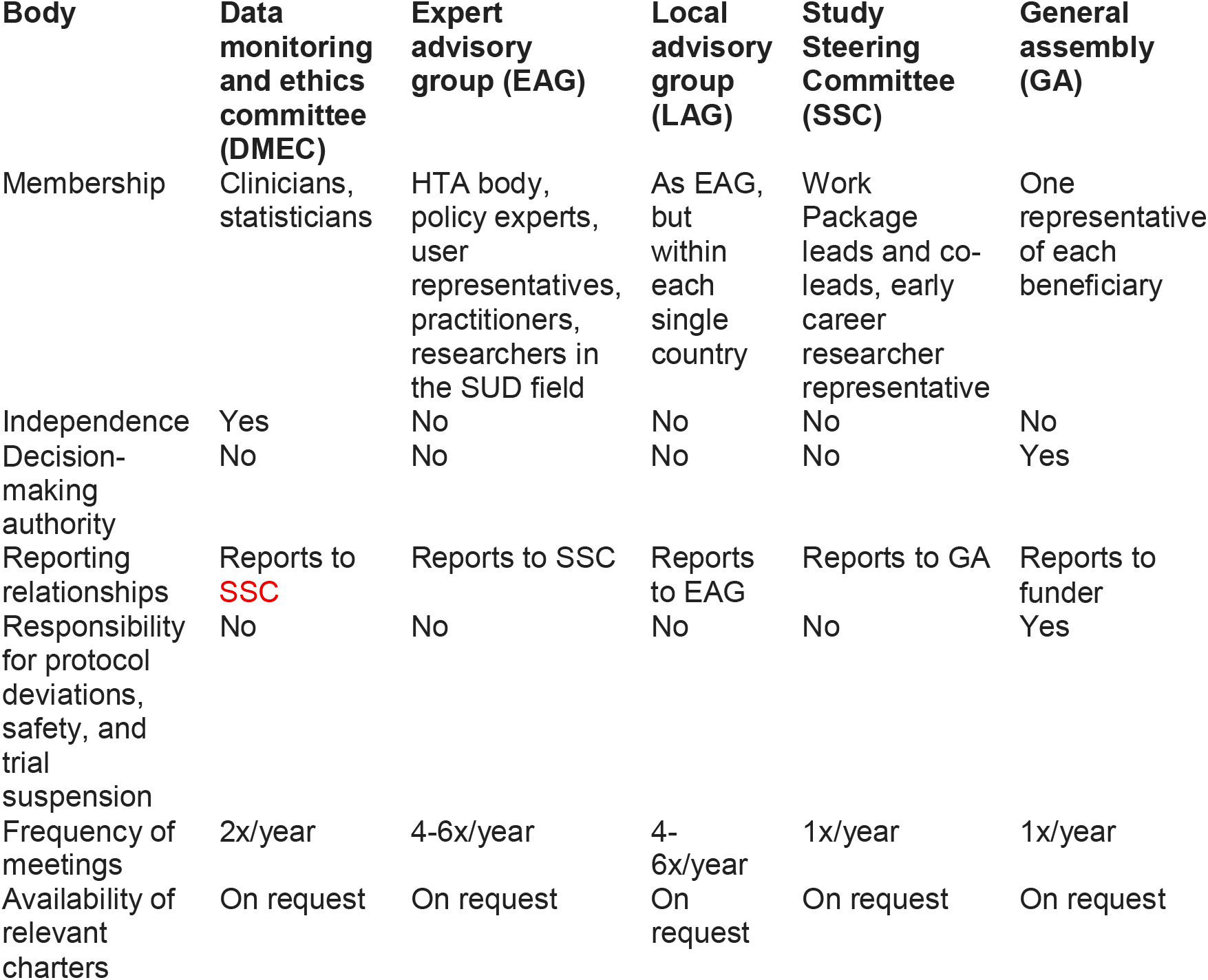
Governance.

## 6. Patient and public involvement

To facilitate user involvement as early in the study as possible, contact with user representatives has been secured through a panel of Norwegian user representatives related to the field of MT (Polyfon Experience Panel). In further preparing the RCT, contact with additional stakeholder organisations in all participating countries and at EU level will be established to ensure optimal connections with future users of the study’s findings. Once the study kicks off, involvement of users will be operationalised at three levels: (1) An Expert Advisory Group (EAG) including a European HTA body (Austrian Institute for Health Technology Assessment, AIHTA), policy experts, user (patients and public) representatives, and experienced practitioners and researchers in the SUD field will be established as an advisory body for the overall study, and to guide all user involvement strategies and activities within the study. The EAG will be organised through a scientific team and include project-employed user representatives and researchers. The scientific team’s responsibility is to secure the scientific methods and processes in the user involvement stages and to interpret the scientific content for the users who are involved and make sure they are familiar with the scientific and methodological framework. To explore and map current practices of user involvement in SUD health care and research contexts across EU countries, and to maximise the study’s impact, the EAG will be involved in reviewing literature, refining methodology, and dissemination of findings, and will assist in ensuring strong links across networks and sister projects. A European-wide association of representatives of intervention services (European Music Therapy Confederation, EMTC) will also be involved to help in recruiting practitioners and to ensure feasibility of interventions and study procedures as well as application of study findings throughout Europe. (2) Local advisory groups (LAGs) comprising user representatives, stakeholders (e.g. policymakers for the health sector, hospital managers) and professionals from the field will respond to the respective local conditions regarding treatment practices, composition of the patients regarding (cultural) background, language, and similar issues. (3) Peer workers will be recruited locally to join professionals at clinical sites in recruitment of patients and administering assessments; they will also act as co-providers of interventions where feasible.

For the user representatives involved in the study, we will aim to achieve similar diversity as among study participants, enabling participants with marginalised identities to be represented by persons with similar backgrounds and thus increase their trust in the treatment and the research process. This way, cultural factors such as various understandings of health and illness can be addressed; personal circumstances like caring obligations that might influence the participation recognised; and different preconditions regarding language skills and literacy considered in the design of consent forms, project information, and other materials. Overall, FALCO’s user involvement activities will contribute with a critical voice within a large RCT. Emphasising the importance of the participants’ own experiences, smaller qualitative studies at different sites (to be described separately) will supplement the findings of the RCT and will give voices to marginalised groups and silenced experiences.

## 7. Ethics and dissemination

This study will be conducted in the declaration of Helsinki. Initial ethical approval was obtained on 2 April, 2025 (Regional Committees for Medical and Health Research Ethics, REK Vest, Norway, reference number 846924) and covers trial procedures in Norway as well as in all other countries. A model consent form is included in the Supplement. In addition, regulations of ethics committees will be followed as applicable in each country. The lead national partner is responsible for following these regulations, and the EU-wide oversight is ensured centrally by NORCE. In each country (Norway, Austria, Israel, Italy, Palestine, Poland, Spain, Switzerland), the lead scientific partner (NORCE, UNIVIE, HERZOG, UNIPV, PNRC, UG, IDIBELL, UZH) will be responsible for preparing and submitting ethics applications according to national rules, including translations and adaptations of patient information and informed consent forms as needed. As this is a non-pharmacological study, regulatory agencies are not involved, and central European ethics applications are not required. Compensation for those who suffer harm from trial participation will be provided according to regulations applicable in each country. Sites have either a general insurance that applies to the trial or have obtained trial-specific insurance. Intervention-related distress will be addressed by interventionists, guided by their supervisors, and clinical teams. Music interventions may be available after the trial if they become a component of the usual treatment at the site. This will depend on the results of the trial and other factors that influence local availability and is outside the scope of the trial.

Protocol amendments will be submitted to relevant ethic bodies, communicated to relevant parties, and recorded on ClinicalTrials.gov. The sponsor is responsible for communicating important amendments to the registry, the main ethics committee, the DMEC, and to site investigators. Site investigators are responsible for communicating them to their local ethics committees, participants, interventionists, and assessors. Interventionists and assessors will also be directly informed by those responsible for intervention and assessments supervision and monitoring.

FALCO will utilise multiple means to share results to further build an understanding of the role of music and music/cultural media/arts-based therapy as a tool in clinical and health contexts, specifically pertaining to SUD, locally and at the EU level, e.g. workshops, meetings, papers/conferences, videos, and infographics. A goal is also to increase knowledge of stakeholders (patients, medical caregivers, researchers, and citizens) on SUD and the role of music interventions, as well as provide a basis for multi-stakeholder collaboration, empowering all to “make and shape decisions” for their health and providing a direct means for increasing health care systems’ benefit from strengthened research and innovation expertise. FALCO will provide local and EU policy makers (clinical health, cultural policy management, health institutions, city government officials) with a range of tools, such as policy briefs, guidelines, comparable contextual models, and specific feedback on the efficacy of music interventions for SUD, specific policy steps for improving their impact, and generally advancing knowledge-based disease management strategies. FALCO will also engage project stakeholders and partners through the organisation of local policy events and an EU Policy Event, increasing clustering and geographical and multidisciplinary synergies.

## 8. Access to data

Anonymized curated data will be made available permanently in publicly accessible repositories using FAIR principles (e.g. using digital object identifiers [DOIs]; open, non-proprietary data formats; clear, consistent meta-data (data dictionaries), using international standards for data structure where available; and licensed for re-use through CC BY). To balance the needs of trial participants for protection of their sensitive health data (in accordance with informed consent) and the needs of the scientific community for data availability, we will follow the principle “as closed as necessary, as open as possible”. Only carefully selected variables will be shared, which do not pose a risk for indirect identification. For variables which may remain potentially indirectly identifiable even after direct identifiers are removed, only the broad category will be shared (e.g. type of substance instead of specific substance). Similarly, shared structural imaging data will only contain the skull-stripped brain scan. With this, we do not anticipate indirectly identifiable data due to small national or clinical subgroups; however, if such subgroups should arise, we will consider relevant measures, such as removing or pooling the respective variable. Informed consent covers explicitly repository deposition and international reuse. Statistical analysis code and data dictionaries will be shared together with the shared data. A statistical analysis plan will be posted at ClinicalTrials.gov before analyses begin. Results summaries will also be posted at ClinicalTrials.gov after the project end.

## 9. Discussion

This manuscript describes the FALCO RCT, the main part of the FALCO project. The overarching goal of the FALCO-project is to examine the effects of MT on different aspects of SUD, reducing disease burden through effective and innovative, yet affordable, treatment. The project will also contribute to the field of MT and SUD-research by elucidating the long-term effects of MT, which are seldom reported on and sorely needed to add knowledge to the field, as well as increase the effectiveness of treatment. Insights gained from this project will form evidence-based recommendations that will inform intervention delivery across Europe and beyond, leading to innovation, safety, and cost-effectiveness in treatment, and improved quality of life for people with SUD. To maximize real-world impact, project stakeholders will actively disseminate results across European countries.

### Strengths and limitations of this study

FALCO is the first large parallel pragmatic RCT to address long-term effects of MT for substance use disorder. Parallel RCTs are the most rigorous method to evaluate comparative effectiveness, and the only appropriate type of RCT when focusing on long-term effects. A drawback, compared to crossover RCTs, is that they require large sample sizes to determine effects.

The broad inclusion criteria, which reflect the pragmatic trial design, will ensure high applicability in practice and will enable examination of subgroups. A drawback is that sufficient test power will be harder to achieve in this pragmatic trial than in an explanatory trial with a more homogeneous sample.

Patient-related stratification variables were considered in dialogue with clinical partners (e.g. opioid maintenance vs. others). Many other clinically relevant variables that were considered may vary quickly over time within individuals (e.g. in-vs. outpatient) or are overlapping (e.g. drug type: alcohol vs. illicit drugs) and may therefore be less meaningful for stratification. That is why we abstained from other stratifications than site.

## 10. Trial status

This is protocol version V1.15, 28 October 2026. History of changes, see Table 1. Start of recruitment was 01 July 2025. The trial is currently recruiting; the first participant was randomised on 26 November 2025. Estimated end of recruitment will be 30 June 2027.

## Supporting information

Supplementary material to the protocol of the FALCO trial

## Data Availability

At the end of the project, anonymized curated data will be made available permanently in publicly accessible repositories (e.g. Open Science Foundation, OSF.io for clinical data and OpenNeuro for brain imaging data).

## List of abbreviations

AIHTA: Austrian Institute for Health Technology Assessment
AMG: Active music group
AUDIT-C: Alcohol Use Disorders Identification Test
CANTAB: Cambridge Neuropsychological Test Automated Battery
COSIMO: COgnition of Social Interaction in MOvies
CV: Cross-validation
DALYs: Disability-adjusted life years
DMEC: Data Monitoring and Ethics Committee
DTI: Diffusion Tensor Imaging
DUDIT-C: Drug Use Disorders Identification Test
EAG: Expert Advisory Group
eBMRQ: extended Barcelona Music Reward Questionnaire
ECR: Early Career Researcher
eCRF: Electronic case report form
EMTC: European Music Therapy Confederation
EQ-5D: EuroQol
EtG: Ethyl glucuronide
EU: European Union
EuropASI: European Addiction Severity Index
FA: Fractional anisotropy
HDBSCAN: Hierarchical Density-Based Spatial Clustering of Applications within Noise
HTA: Health Technology Assessment
HUMS: Healthy-Unhealthy Music Scale
GA: General Assembly
GBDT: Gradient Boosted Decision Tree
GFAP: Glial fibrillary acidic protein
ICD-10: International Classification of Diseases, Tenth Revision
ICR: Importance, Confidence, Readiness Ruler
ID/ED: Intra-Dimensional/Extra-Dimensional Set-Shifting
IQ: Intelligence Quotient
LAG: Local advisory group
LC-MS/MS: Liquid Chromatography-Tandem Mass Spectrometry
MADRS: Montgomery-Åsberg Depression Rating Scale
MAE: Mean Absolute Error
MASC: Movie for the Assessment of Social Cognition
MBI: Mindfulness-Based Intervention
MDMA: Methylenedioxymethamphetamine
MI: Motivational Interviewing
ML: Machine Learning
MLG: Music listening group
MRI: Magnetic Resonance Imaging
MT: Music therapy
NfL: Neurofilament light chain
PAL: Paired Associates Learning
PI: Principal Investigator
RD: Radial diffusivity
PROMS: Profile of Music Perception Skills
RCT: Randomised Controlled Trial
RVP: Rapid Visual Information Processing
SAP: Statistical Analysis Plan
SCI: Sleep Condition Indicator
SHAP: SHapley Additive exPlanations
SHAPS: Snaith-Hamilton Pleasure Scale
SM: Supplementary Material
SSC: Study Steering Committee
SUD: Substance Use Disorder
SURE: Substance Use Recovery Evaluator
SWAT: Study within a trial
SWM: Spatial Working Memory
TAU: Treatment as usual
TCIA: Test of Creative Imagery Abilities
THC: Tetrahydrocannabinol
t-SNE: t-distributed Stochastic Neighbour Embedding
URICA: University of Rhode Island Change Assessment
VBM: voxel-based morphometry
WAIS-IV: Wechsler Adult Intelligence Scale 4th Version
WP: Work Package
2C-B: 4-bromo-2,5-dimethoxyphenethylamine

## Declarations

### Ethics approval and consent to participate

This project has been approved by the Regional Committees for Medical and Health Research Ethics (REK), Region West (846924). Ethics approval will also be obtained from the responsible ethics committees in each country prior to recruitment. The project has been registered at ClinicalTrials.gov (NCT 07028983). All participants deemed eligible will receive extensive information about data collected during the study and will have to provide written consent before participation.

### Consent for publication

Not applicable.

### Availability of data and materials

At the end of the project, anonymized curated data will be made available permanently in publicly accessible repositories (e.g. Open Science Foundation, OSF.io for clinical data and OpenNeuro for brain imaging data) using FAIR principles (e.g. using digital object identifiers [DOIs]; open, non-proprietary data formats; clear, consistent meta-data, using international standards for data structure where available; and licensed for re-use through CC BY).

### Authors’ contributions

MG, JA, KD, BJ, SK, AM, MSo, LT, AHE, MP, FS, GS, DS, ARF, ES, AVB, MH, BBQ, and CG conceptualised the ideas and formulated or developed the overarching research goals and aims.

MG, JA, SK, LT, AHE, TSS, LL, MP, FS, GS, DS, EC, VFD, ARF, ES, AVB, MH, BBQ, and CG developed design of methodology and created models.

GS, ARF, and DV provided resources in the form of study materials, reagents, materials, patients, laboratory samples, instrumentation, computing resources or other analysis tools.

AUK, MSt, and FM contributed with data curation by producing metadata and maintaining research data code for initial and later reuse.

CG wrote the initial draft. BK contributed to writing the sections background and participants; MG, JF, TG, interventions; AHE, long-term outcomes; BBQ, biological samples; AHE, AVB, BBQ, neuropsychological tests; SK, ARF, MH, BBQ, structural brain imaging; JA, sample size and statistical analyses; FS, DS, machine learning analyses; MG, LT, JF, patient and public involvement.

All authors provided review, commentary and revision of the draft.

MG, AHE, BK, and CG contributed to visualisation by creating or presenting published work and data presentation. CG created Table 1 and Figures 1-5. AHE and BK created Table 2. MG created Table 3.

MG, SK, AHE, LL, MP, FS, GS, FM, TS, IZK, TG, MB, LFP, PP, ŁB, EC, VFD, ARF, AVB, SFM, DV, MH, BBQ, and CG had leadership responsibility for the planning and execution of research activity and had the role of supervisor.

MG, HMKM, AS, CLD, AHE, JH, BK, LL, HAR, FS, GS, FM, ŁB, ES, MH, and BBQ contributed with project administration by managing and coordinating research activity planning and execution.

MG, CLD, AHE, LL, MP, FS, ARF, AVB, DV, MH, BBQ, and CG acquired financial support for the project.

## Acknowledgements

We would like to thank Paul Bassett, Xi-Jing Chen, Geert Dom, Jörg Fachner, and Michael J. Silverman for serving as members of the Data Monitoring and Ethics Committee and for providing valuable input.

