## Supplementary material to the protocol of the FALCO trial for "Fighting Addictions, improving Lives through COmprehensive rehabilitation with music (FALCO): Protocol for an international randomised controlled trial"

### 1. Text of model consent form (Version 5, December 2025)

DO YOU WANT TO PARTICIPATE IN THE RESEARCH PROJECT "FIGHTING ADDICTIONS, IMPROVING LIVES: COMPREHENSIVE DRUG REHABILITATION WITH MUSIC (FALCO)”?

PURPOSE OF THE PROJECT AND WHY YOU ARE INVITED TO PARTICIPATE

You are being invited to take part in this project because you are currently in contact with [insert name of institution]. In this research project, we aim to investigate how music therapy can contribute to improve the treatment of substance use disorder (SUD).

Background for the project

Music is a part of our lives, both in everyday situations and during significant moments. Research shows that music therapy can contribute to improve substance use recovery, but we know little about whether music therapy has lasting effects on addiction problems. This project will investigate whether combining music therapy with standard treatment contributes to improved treatment of substance use disorders long-term.

How the study works

This study is led by NORCE Research AS (Bergen, Norway) in collaboration with several research organizations in Norway and internationally (please find a complete, updated list on the project website, www.falco-project.eu). The Principal Investigator is Professor Christian Gold at NORCE.

In Norway, a total of 180 individuals will take part in the study, and participation will take two years. The same study will be carried out in parallel in seven countries, with a total of 600 participants.

Participation is free and voluntary. NORCE has received research funding from the EU’s HORIZON research program to carry out this project. We are also working with several user organizations and national and international experts to ensure the study is relevant and of high quality.

WHAT PARTICIPATION IN THE PROJECT INVOLVES FOR YOU

By participating in this study, you will help build important knowledge about how to treat substance use disorders – knowledge that could greatly benefit society. The results may support the use of group-based music therapy as an effective part of addiction rehabilitation.

Participation timeline

Participation in the study means that you will be asked to follow an assigned treatment program for 1 year and complete health assessments over a 2-year period (including any optional continued treatment during the second year). During this 2-year period, you will complete assessments of various aspects of your health, including substance use, mental health, cognitive functioning (such as your ability to think, learn, and remember), motivation, and quality of life. These assessments will be conducted at the start of the project and repeated after 3 months, 1 year, and 2 years. Each assessment includes set of questionnaires and behavioral tests (required), biological samples (optional), and brain imaging (optional, and only available to participants in Bergen). The first assessment will take about 5 hours (which can be split over two days), while follow-up assessment will take about 3 hours. The total time these assessments will take will vary depending on whether you choose to contribute with biological samples and brain imaging.

The treatment program:

Those who agree to participate in the study, will be randomly assigned to one of three treatment groups. Each group is equally important for helping us understand whether music therapy has an impact on treatment outcomes. Therefore, all participants will be randomly allocated to one of the following three groups through an anonymous random selection:

•	Standard Treatment without Music Therapy: In this group, participants will continue with regular treatment during the first year. Participants selected to this group will be offered music therapy after the first 12 months of the study (during the second year).

•	Standard Treatment with Music Listening Therapy: In this group, participants get the opportunity to listen to music, reflect on the role of music in their lives, analyze lyrics, and take part in group discussions about these experiences.

•	Standard Treatment with Active Music Therapy: In this group, participants will get the opportunity to play musical instruments, improvise, compose music, write songs, and more together with other group members. You do not need any musical experience – no singing or playing skills are required.

If you consent to participate and are assigned to a group, you are expected to attend once or twice a week, depending on your standard treatment and whether you were assigned to a group with music therapy. If you are assigned to a music therapy group, you will be expected to continue your standard treatment as arranged with your treatment provider and to attend weekly music therapy sessions. Music therapy sessions will last about 90 minutes, once a week, on a fixed day and time agreed upon with the music therapist and the other group members. Treatment will continue for 12 months. As a part of the research, we will make video recordings of the music therapy sessions to study how they are carried out. Should we see a need to use selected recordings in a wider context (for educational purposes or at conferences), we will ask your consent for this separately.

Questionnaires and behavioral tests (relevant for all participants):

We will collect information about your health using various questionnaires where you will be asked about aspects such as your age, education, diagnoses, substance use, quality of life, mental health, sleep, and daily functioning. In addition, we will gather information about your cognitive functioning. These assessments will take place at the start of the project and will be repeated after 3 months, 1 year, and 2 years. After two years, your participation will be concluded. After this, we will also collect data from public records to map your employment, any hospitalizations, any convictions, and survival after ten years, but you do not need to contribute to this part of the data collection.

Biological sample tests (relevant for participants who are eligible):

Biological samples will be collected at the start of the project and will be repeated after 3 months and 1 year. Hair samples will be collected by taking three strands of hair (3 cm long) from your scalp (or possibly an alternative place). The samples will be collected by personnel trained in hair sampling. You will also complete a brief questionnaire. We will collect 10 ml of venous blood, drawn by trained medical staff. These samples will be analyzed for biological markers related to brain and neural health, such as neurofilament light chain (NfL), glial fibrillary acidic protein (GFAP), and phosphorylated tau (p-tau), which serve as indicators of brain stress associated with substance use.

Brain imaging (relevant for participants who are eligible):

We will measure effects on brain structure and function using magnetic resonance imaging (MRI). MRI is a medical imaging technique that does not use harmful radiation. Instead, it uses radio waves generated within a magnetic field inside an MRI scanner. When these waves interact with different tissues in the body, a radio echo is sent back and captured by a small antenna in the machine. These signals are then turned into digital images. By using various techniques, we can, for example, measure blood flow in the brain to gain insight into brain activity—this use of MRI is called functional MRI, or fMRI. We will also use the MRI scanner to perform Diffusion Tensor Imaging (DTI), which produces digital images used to visualize and map the nerve fibers that connect different parts of the brain. In this part of the study, we use fMRI to examine how the brain responds to drug-related and neutral images, combined with different types of music. The goal is to better understand how music therapy affects the brain’s reward system and how this may help reduce cravings for substances.

The examination involves you lying on your back on a bench that slides into the MRI scanner. You will be asked to stay completely still during the scan. The MRI scanner makes loud noises, but you will be given ear protection to reduce discomfort and protect your hearing. During the scan, you should lie as still as possible, and you will be asked to look at images and press buttons to tell us how you react to them. During the scan, you will look at images and listen to music through headphones. The images will either show drug-related situations (such as alcohol or other substances you have used) or neutral images. At the same time, you will listen to either pleasant or unpleasant music. After every sequence, you will be asked to evaluate how much you crave the substance and how pleasant you found the music to be.

The MRI scans will take place in the radiology department at Haukeland University Hospital, in the basement of Sentralblokken (MRU3). Scans will be done at the beginning of the study, and again after 3 and 12 months. After each completed MRI session, you will receive NOK 200 as compensation for your time.

POSSIBLE BENEFITS AND RISKS

Participation in this study will not affect your standard treatment. By participating, you have the opportunity to receive music therapy, which is intended to help you manage challenges related to substance use. Whether or not this therapy is effective is part of what our research aims to answer. The results of the study will serve as an important foundation for further research and the development of therapeutic methods. After the study ends, you will receive detailed information about the results and publications from the project.

Some topics discussed in the music-focused group sessions may bring up strong emotions, thoughts, and experiences that could be perceived as distressing (such as difficulties participating in groups, lack of motivation for rehabilitation, challenges in managing addiction, shame, anxiety, etc.). These feelings can be shared within the music therapy group, where music therapists will be able to support you throughout the process.

Participants assigned to the standard treatment without music therapy may feel disadvantaged by not being able to participate in music therapy initially. However, you will be offered music therapy after 12 months.

Biological samples:

Hair samples pose no health risk and have only minimal cosmetic impact. A very small amount of hair is cut—so little that it is usually not noticeable. The collection process takes less than five minutes to complete.

Risks associated with blood sampling include the possibility of developing a small bruise, redness and/or swelling at the puncture site, as well as a local infection. However, when the procedure is performed correctly, these risks are very low.

Brain imaging:

MRI examinations are generally without complications. As far as we know, there are no known risks or side effects from MRI (fMRI) scans. However, it is important to follow the safety instructions provided before and during the examination. Because of the strong magnetic field of the MRI machine (thousands of times stronger than the Earth's magnetic field), people cannot participate if they have metal objects and/or pacemakers implanted in their bodies. People with fixed dental implants can usually be scanned safely. For other metal implants (such as artificial joints, metal plates from bone fractures, etc.), the radiographers operating the MRI machine and the study coordinators will assess on a case-by-case basis, whether you can undergo the examination. It is important that you speak up if you are unsure or have any questions. During the scan, you will be inside a relatively narrow tube, which may feel uncomfortable for people with claustrophobia. However, you can call for assistance at any time during the scan and communicate with the radiographers operating the MRI machine using an intercom system. You may stop the examination at any point if you feel uncomfortable.

The images used in this part of the study may bring back memories or trigger cravings. This is expected and necessary to measure how music therapy affects these kinds of reactions in the brain. Therefore, we have several safety measures in place:

•	You will be informed in advance about the content of the images.

•	You can stop the examination at any time without having to give a reason.

•	You will be in contact with the research team at all times during the scan.

•	After the scan, you will have a thorough conversation with a researcher who will help you to make sense of the experience in relation to your treatment.

•	If you wish, your clinical contact person can be involved right away.

•	There will be no direct access to any drugs during or after the examination, and it will all take place in a safe clinical setting.

What happens if something unexpected shows up in the MRI scans?

The MRI scans are done for research purposes and are not meant to be clinical examinations. However, a radiologist will review the images. If something is found that might be medically important, a doctor from the project will contact you and refer you for follow-up with your general practitioner or a specialist.

VOLUNTARY PARTICIPATION AND YOUR RIGHT TO WITHDRAW CONSENT

Participation in the project is completely voluntary. If you choose to participate, you will be asked to sign the consent form on the final page of this letter. You can withdraw your consent at any time, without giving a reason. If you choose not to participate or decide to participate and withdraw later, it will not have any negative consequences for you or your standard treatment.

If you withdraw your consent, your data will no longer be used for research purposes. You have the right to access the information stored about you, which will be provided within 30 days upon request. You can also request that your data be deleted from the project. However, the right to request destruction, deletion, or disclosure does not apply if the material or data has already been anonymized or published. These rights may also be limited if the data has already been included in completed analyses. If you have questions about the project or wish to withdraw later, you can contact your therapist, the project coordinator, or project leader (see contact information on the final page).

The principal investigator may also, at their discretion, withdraw you from the study if: a) there is a change in your health status that can affect the study, b) you no longer meet the minimum requirements for participation, or c) the study is discontinued for any reason.

WHAT HAPPENS TO YOUR DATA

The information collected about you will only be used as described in the purpose of the project and is planned to be used until 2039. Any extension of use or storage period can only occur with approval from the Regional Committee for Medical and Health Research Ethics (REK) and other relevant authorities. You have the right to access the information registered about you and to have any errors corrected. You also have the right to access information about the security measures in place for handling your data. You may file a complaint about the processing of your data with the Norwegian Data Protection Authority (Datatilsynet) and the institution’s Data Protection Officer.

All data will be processed without your name, personal ID number, or other directly identifying information – this is called coded data. A code links you to your data through a list containing names and personal ID numbers, and only authorized personnel associated with the project will have access to this list.

Publishing results is a necessary part of the research process. All publications will be made in a way that prevents individual participants from being identified. However, we are required to inform you that complete anonymity cannot be absolutely guaranteed, though the risk of being identified is very low. Your information will not be shared with the authorities, including the police.

Once the study is complete, the information about you (the list with names and personal ID numbers) will be stored for five years for auditing purposes. After that, this information will be permanently deleted.

DATA SHARING AND TRANSFER ABROAD

By participating in this study, you also consent to making coded information about you (such as age, gender, data from questionnaires and behavioral tests, MRI data, and biological samples) available to international research partners as part of collaborative research in line with the project’s stated purpose described earlier. Collected data will be made available via SAFE (the University of Bergen’s secure server for research data), where collaboration partners receive remote access without downloading. The transfer is carried out in accordance with European data protection legislation (GDPR), and the code linking you to your personally identifiable information (name, personal ID number) will not be shared.

NORCE is responsible for collecting and sharing information about you. All collaborating partners enter into an agreement that the data will only be used for the purposes of the project. Each collaborating partner has its own responsibility for the use of the data. More information about this agreement and the use of your data in each work package is available on the project’s website, www.falco-project.eu.

After the project ends, anonymized research data (such as clinical and brain imaging data, not your name or personal ID number) will be made openly available for reuse by other researchers in secure international data repositories (such as Open Science Foundation (OSF.io), Frankfurt, Germany for clinical data and OpenNeuro for brain imaging data) for an indefinite period. This ensures that the research findings can be verified by others, which is important for the credibility of the research results.

WHAT HAPPENS TO THE SAMPLES TAKEN FROM YOU

The biological samples you provide will be stored in a research biobank associated with this project. The biobank is located with a project partner in Switzerland, who will be responsible for analyzing the samples. The biobank will be closed at the end of the project, and all materials will be destroyed.

INSURANCE

NORCE holds specific corporate and product liability insurance, professional liability insurance, and occupational injury insurance. For interventions carried out by healthcare personnel within a healthcare institution, the Norwegian Patient Injury Compensation scheme will cover any potential harm resulting from participation in the study.

FOLLOW-UP PROJECTS

If a follow-up project becomes relevant, we will contact you.

APPROVALS

Regional Committee for Medical and Health Research Ethics (REK) has conducted an ethical review and approved the project (REK #846924).

We process your personal information for research purposes in the public interest, and participation is entirely voluntary.

CONTACT INFORMATION

If you have questions about the project or wish to withdraw your participation, you can contact your therapist or contact:

If you have any questions about data protection in the project, you can contact NORCE’s Data Protection Officer at:

I CONSENT TO PARTICIPATE IN THE PROJECT AND TO THE USE OF MY PERSONAL DATA AS DESCRIBED

•	I have read and understand the information about the study.

•	I have been informed about possible benefits and risks of participating in the study.

•	I have been informed about how my information will be processed.

•	I am aware that I will only be compensated for MRI examinations I participate in.

•	I am aware that I can withdraw from the study at any time without having to provide an explanation and that there will be no negative consequences to my standard treatment if I choose not to participate or withdraw from the study at a later time.

☐Yes	☐No	I consent to participate in the collection of hair samples.

☐Yes	☐No	I consent to participate in the collection of blood samples.

☐Yes	☐No	I consent to participate in the collection of brain images.

Place and date	Participant’s signature

Participant’s name in block letters

I confirm having given information about the project

---------------------------------------------------------------------------------------------------------------------------------------------------

Place and date	Signature	Project role

### 2. List of interventions recorded as part of TAU

***Specific medications for opioids abuse***

Buprenorphine

Methadone

Naltrexone

Suboxone (Buprenorphine+Naltrexone)

Naloxone

***Specific medications for the treatment of alcohol use disorders***

Disulfiram

Acamprosate

GHB

Selegiline

GHB / Sodium oxybate

Baclofen

Pregabalin

***Specific medications for the treatment of nicotine use disorder***

Yes

No

***Psychopharmacological therapies for psychiatric comorbidities***

Antipsychotics

Antidepressants

Benzodiazepines

Mood stabilizers/anticonvulsants

ADHD medication

***Psychosocial treatment***

Cognitive behavioral therapy

Dialectical behavioral therapy

Mentalization-based therapy

Psychoanalysis

Mindfulness-based therapy

Cognitive remediation

Motivational interview

Mutual help/self-help

Family therapy

Contingency management

Structured physical activity

Occupational and art-based therapies

***Frequency of each psychosocial treatment***

Once per week or more

Less than once per week

### 3. Text of intervention manual (version 1.2, May 2026)

1.	Introduction

The following intervention guidelines have been created for the implementation of two group music therapy settings, AMG (Active Music Group) and MLG (Music Listening Group), to be provided to participants with substance use disorder (SUD) in the FALCO project. They have been developed based on available literature on music therapy in the context of SUD, and informed by further articles describing the development of music intervention manuals for certain populations. Principles, goals and intervention elements described in these guidelines were then formulated in a series of discussion meetings among the authors involving their own clinical experience as music therapists.

Interventionists need to be qualified music therapists with a Master’s degree, with a minimum of one year of full-time equivalent experience working as a music therapist in a group setting with individuals with SUD. Before implementing sessions in FALCO, therapists will undergo a study-specific training to be familiar with and competent in applying these intervention guidelines, and in following general study procedures including videorecording of sessions and documentation. Furthermore, regular online supervision in English for all interventionists who are involved in FALCO will be provided throughout the study to ensure tailored application of the guidelines, and to provide support to the interventionists. Where feasible, peer workers (individuals with lived experience of a SUD who have undergone specific training to offer support to people with SUD alongside professionals) will act as co-providers of interventions. They will also undergo study-specific training workshops and supervision.

Providing sets of basic principles (section 3), goals (section 4), and intervention elements (section 5), these guidelines are intended to work independently of interventionists’ specific training backgrounds and theoretical orientations. Based on their own clinical judgment, therapists need to apply them flexibly, focusing on goals and choosing activities based on individual participants’ and the group’s needs, cultural contexts, and local circumstances.

2.	General comments regarding the setting

2.1.	Group size

Groups in FALCO comprise around 6 participants. Groups should ideally start with no less than 4 participants. Depending on how recruitment is organised at each site, groups may be closed (with no participants joining after its start) or open (where it is possible that participants join the group during its duration, and participants might leave the group if they have completed their 12-month participation or if they discontinue participation for any other reason). The number of participants per group should not exceed 10 at any time to allow for enough time and space for participants within each session; the maximum number also depends on the size and equipment of the room available at each clinical site (see D5.1 checklist). Group sessions may be supplemented with individual sessions if needed (e.g., to prepare for groups).

2.2.	Session duration

Each music group session should last between 60 and 90 minutes (depending on local circumstances and participants’ needs), with additional 15 minutes pre-session for the therapist to prepare the session and for arrival of and informal exchange between participants (possibly with refreshments such as tea in cold weather or cold water during summer time) and 15 minutes post-session for the therapist to clear up the room and fill in study-related documentation forms, and for further informal exchange between and departure of the participants.

2.3.	Safe setting

To establish a respectful environment and a sufficiently safe space for everyone, remind the participants to:

(a)	maintain respectful language and attitude towards each other throughout the pro-cess, and emphasise that in case of misunderstandings or conflicts, it is important to communicate efficiently and respectfully;

(b)	keep confidentiality;

(c)	have phones, smart watches and similar devices switched off or in silent mode and/or left in a dedicated space (e.g., a box) during sessions.

(d)	Agree on crisis strategies for cases of overwhelming experiences and emotional dis-tress (e.g., temporarily leave the room to get support from another professional if available; provide helpline numbers or direct therapist contacts for emergencies).

(e)	Discuss within the group if members wish for any further group rules, expectations, or individual aspects to consider (e.g., fears, concerns, and things or music to avoid).

Whenever a new member joins the group, make sure to inform them about these requirements to ensure a sufficiently safe setting.

If the institution you work in has specific rules on conditions in which participants cannot join group sessions (e.g. if they are intoxicated or currently consuming), these also apply to sessions provided as part of FALCO. If there are no specific rules at the institution you work in, we strongly recommend establishing adequate regulations on any conditions and related procedures within your local team to ensure sufficient safety for all involved. Once agreed upon locally, make sure to clearly communicate such additional rules to the participants.

2.4.	Provision, attendance & fidelity

Sessions will be offered to participants once a week for at least one year. Where feasible, a peer worker (expert by experience with pertinent training) will co-provide sessions. Groups will be assembled by clinical site managers or therapists based on clinical, personal, and practical aspects (e.g., substance type, stage in the recovery process, clinical setting type, scheduling).

Participants’ attendance of AMG/MLG sessions will be captured by entering the dates of the first session, the number of sessions attended since randomisation, and the date of the final session in REDCap. After each session, a self-report form and checklist (currently in development, see below in sections 4 and 5 and Appendix) will be completed by the therapist so that documentation of provision and participants’ attendance of sessions as well as treatment fidelity is documented.

For evaluation of treatment fidelity by independent assessors, sessions will be videorecorded wherever feasible using a smartphone, tablet or laptop, with video files streamed directly during recording via a secure web service (“Sikker Video”) and stored centrally on a safe desktop called SAFE. These IT structures are provided by the University of Bergen (UIB) and ensure that the video data, including sensitive personal information, are transmitted and stored securely. A detailed guide on how to set up Sikker Video and access to SAFE will be available separately (Guide for GAMUT SAFE provo and sikker-video v3.docx).

3.	Basic principles

3.1.	Sessions follow a general structure to provide predictability

It is recommended that each session includes three main parts, to foster predictability and stability, and to ensure consistency of intervention delivery across therapists and sites:

(1) Opening warm-up (musical and/or verbal); (2) music experience(s), with space for verbal reflection; (3) a closing round to reflect upon the group experience. The exact duration of each of these parts will depend on participants’ needs and the therapist’s clinical judgment and can also be determined by active involvement of participants. It is also recommended to provide as much consistency as possible in temporal and spatial features of the group (e.g. regarding day of the week, room for sessions).

3.2.	Exploration, curiosity, not-knowing stance

To foster participants’ agency and co-construction of meaning, the overall stance of the therapist is grounded in active listening, modelling curiosity and interest, and conveying a stance of “not-knowing” (i.e., to refrain from expressing assumptions about the participants’ lived experience), thereby facilitating a dynamic interplay between expertise, reflexivity, and ongoing learning among all members of the group.

3.3.	Regulating group phases

The therapist’s style of group leadership changes depending on the developmental level of the group as well as setting conditions (e.g. timeframes), with more guidance and a higher level of structure at the beginning and end of the group process as well as each session. It is recommended that the therapist promotes group engagement with music quickly and early on in the therapeutic process. When approaching the end of each group member’s phase of participation (i.e., usually one year after randomisation), the therapist together with group members addresses this appropriately both psychologically and musically (e.g., during the last month, discuss ways to conclude participation, ‘say/play goodbye’, or produce ‘take-away’ items such as playlists, group recordings, lyrics notebooks etc.).

3.4.	Active promotion of group cohesion

The therapist works actively to promote a sense of trust, group wholeness and belonging through music activities and discussion, especially in early stages of the group (foster member-to-member interaction, encourage to learn from each other, feedback to the group, etc.). While supporting group cohesion, the therapist simultaneously addresses individuals’ and sub-groups’ needs as required.

3.5.	Integrate use of music inside and outside of sessions

The therapist addresses and explores connections between music, group experiences and everyday life, with a special focus on individually beneficial and harmful uses of music. Throughout activities, it is paramount to raise awareness regarding feelings, sensations (e.g. craving) and behaviours (e.g. using substances) that may be provoked during music experiences, including awareness of risky uses of music within and outside of sessions. Throughout the process, the therapist supports participants to develop strategies to manage music experiences without risking relapse or other harm. This includes learning to avoid certain uses of music if necessary, finding and using alternative favourite music types, and ultimately developing tolerance to music experiences previously associated with substance use. The music therapist supports the participants’ use of music in everyday life as a self-care method and explores together with participants any changes in their music use that might occur between sessions as a result of their experiences and new insights gained.

4.	Goals

The following goals are based on a survey of the literature combined with clinical and ser-vice user expertise and experience. The goals represent core achievements that partici-pants with SUD might aim to achieve through music interventions. The order of the goals is not supposed to indicate a certain hierarchy; the relevance and importance of goals may vary in regard to each participant, group, and phase of intervention.

Each goal may be pursued by and focused on through several of the intervention elements/music experiences/musical activities described in section 5, with the possibility of facilitating an experience that may serve several of these goals at the same time. For assessment of treatment fidelity and intervention progress over time, therapists will be asked to indicate intervention elements implemented and goals that were discussed/aimed at during the sessions in specifically developed documentation forms including a table that connects goals with music activities (see Appendix).

5.	Intervention elements / music experiences / musical activities

In this section, suggestions for musical activities to conduct in either the Active Music Group or the Music Listening Group are presented. They are meant to be applicable independently of the specific music therapy training backgrounds of the therapists involved. However, specific activities may be more prevalent in one music therapy tradition than another, and so therapists should only offer those activities which they feel confident in delivering. Therapists are invited to contact the working group members in case they need more context or details on some of the activities.

5.1.	Active music group (AMG)

The focus of music experiences in this intervention type is on active music-making.

5.1.1.	Composing music (instrumental or vocal, no lyrics)

•	Choose a topic/story OR invite the participants to choose a topic/story for the musi-cal piece (e.g., relating to daily life, identity, emotional states).

•	Invite participants to concretise the topics and describe details.

•	Guide the participants’ discussion on aesthetic qualities, styles, sounds, and in-struments that might be appropriate.

•	Concretise the participants’ suggestions into musical features such as time, rhythm, harmony, and melody; assist as much as necessary to turn their ideas into sonic re-ality and finalise the composition.

•	Play/sing the composition with the chosen instruments/electronic devices.

•	Write down or record the piece; if recorded, decide together with the group mem-bers where recordings will be stored and shared.

•	This activity can be implemented across several sessions, depending on the process of developing the composition.

5.1.2.	Songwriting

•	Choose a topic OR invite the participants to choose a topic (e.g., relating to daily life, identity, emotional states).

•	Suggest an appropriate style/musical framework/melody for the song, and play to the participants (alternatively, an already existing song matching with the topic or the participants’ musical preferences might be chosen and listened to).

•	Invite participants to brainstorm according to the topic (interchangeable with the latter).

•	Compile ideas for lyrics together with the participants, write down lyrics and melody (if applicable).

•	Sing the song, invite participants to join in, and accompany on an instrument.

•	If desired, the song might be recorded; if so, decide together with the group members where recordings will be stored and shared.

5.1.3.	Improvisation: free

•	Invite participants to try out different instruments in the room (including body per-cussion, voice, and movement), and eventually choose one or more instruments.

•	Encourage the participants to play a short phrase on their chosen instrument (all to-gether or taking turns).

•	Start playing a simple harmonic frame (on a piano or guitar) OR a steady beat (on a well-resonating drum, such as a djembe) and encourage the participants to join in OR let the improvisation evolve by itself.

•	Adjust your playing to the group needs/dynamics (e.g., providing a steady ground, withdrawing, dialoguing, etc.).

•	Let the improvisation end ‘naturally’ without your guidance OR let the participants end it with some signal from their side OR guide it to an end musically or verbally if participants do not end the improvisation themselves.

•	Encourage the participants to share their experiences with the group and offer your observations/impressions.

•	Highlight themes that were expressed by the participants and frame them in words.

5.1.4.	Improvisation: (pre-)structured

Modes of (pre-)structuring an improvisation, which can also be combined and serve as supplements to the overall structure of an improvisation as described above, include:

-	Pre-structuring by musical parameters: Agree in advance on progressions like soft – loud – soft, slow – fast – slow.

-	Pre-structuring by a theme/topic: Suggest a theme OR encourage participants to suggest themes to serve as a title/topic for the improvisation in advance, find agreement on one title/topic, and begin improvising accordingly. Themes can focus on various emotional states, especially ones which are being avoided or dampened through substance use, where expressing them musically can serve as practice to cope with them without using substances.

-	Pre-structuring by a certain (group of) instrument(s) to be played exclusively: Invite participants to choose one instrument out of the stockpile of percussion OR melod-ic/mixed instruments OR use their voices exclusively. Invite participants to briefly explore the instruments before starting to improvise together.

-	Distributing roles: Agree on participants to take over certain roles during the im-provisation. Examples are: conducting the group’s playing by hand signs of forte/piano, slow/fast, stop/start, etc.; leading during the improvisation, e.g., by set-ting the tempo, volume, etc. directly in the music; having group members play/sing for one participant exclusively, e.g. to musically explore a specific topic or to receive a ‘sound bath’; have two participants play together with others listening and observ-ing their own reactions.

-	Structuring musically: Guide the group through your play, e.g., gradual transitions – crescendo/diminuendo, piano/forte, soft sounds/shrill sounds, etc.

5.1.5.	Educational activities: teach basic musical skills

•	Focus on certain basic playing skills/rhythms/melodic and rhythmic structures.

•	Choose instrument/s (e.g., drums, percussion, band instruments, piano, body per-cussion, movement/dance).

•	Show participants basic skills on the chosen instruments (e.g., chords, playing modes).

•	Choose teaching mode using visual cues or notation (e.g., cards with simple signs for notation, tempo).

•	Start with a basic rhythm (e.g., rock, pop, bossa nova), which participants gradually complement with additional rhythms OR with basic chords to gradually complement with melody OR teach and initiate elements like “call and response”, etc.

•	Gather feedback and discuss experiences.

5.1.6.	Educational activities: enable participants to play as a band or sing as a choir

•	Choose song/piece OR encourage participants to choose songs/pieces for the joint singing or band playing.

•	Distribute roles.

•	Organise learning in partial-group and whole-group setting.

If considered feasible and desired by the participants to work towards a concert:

•	Choose setting in which a performance is possible (i.e., location, context, room, au-dience, time of day, date).

•	Choose song/s or piece/s OR encourage participants to choose songs/pieces for the performance.

•	Decide on schedule for rehearsing within group sessions.

•	Manage motivational aspects and reflect on social interaction within the group dur-ing rehearsing period.

•	Decide together with the group members if recording will take place, and if so, where recordings will be stored and shared.

•	Decide together with the group what happens immediately after the performance and in subsequent sessions.

5.2.	Music listening group (MLG)

The focus of music experiences in this intervention type is on listening to music, without active music-making or use of other artistic expression (such as drawing).

For all music listening activities, the therapist makes sure to either propose or invite participants to choose a suitable position during listening – sitting, lying down (if adequate mats are available), moving around in the room, etc.

5.2.1.	Music listening with a focus on music in everyday life

•	Invite participants to discuss their everyday use of music.

•	Raise awareness to how they feel after listening to or playing different musical piec-es and discuss the variety of feelings evoked.

•	To address non-beneficial uses of music, and if in the appropriate group phase (i.e., if the level of safety and mutual trust is judged as sufficient), invite participants to choose a piece of music that is connected to their substance use (and might provoke craving and/or relapse), listen to it in the group and discuss the feelings related to it.

•	Support participants in forming new associations with this music (particularly those related to recovery and rehabilitation) and/or in avoiding certain uses of music when necessary.

•	Additionally, help them identify alternative music that promotes enjoyment without being linked to substance use, and that can divert their mind from negative thoughts and images, especially when experiencing craving.

•	Based on this, developing individual playlists for specific purposes might be a way for participants to integrate beneficial uses of music in everyday life.

5.2.2.	Music listening with a focus on identity / life stories

•	Encourage a participant to select (either spontaneously or in preparation for an up-coming session) one or more pieces of music and sounds that represent “who I am” OR “how my life has been so far” OR “three pieces of music representing my past, present and future”.

•	Guide the group members to pay attention to two aspects: a) how the music affects them; b) what they learn about the presenter through the music.

•	Listen to the music in the group.

•	Ask the presenter whether they prefer to first explain why they chose this music and then hear what the group members have to say, or the opposite.

•	Encourage a respectful discussion around the music between group members.

5.2.3.	Music listening with a focus on regulation of tension and emotions

•	Choose a musical piece/instrument/sound that can support regulation of tension and emotions OR invite the participants to choose.

•	Invite the participants to find a comfortable position that allows for relaxation.

•	Encourage participants to observe their level of tension and any emotions immedi-ately before listening.

•	Play pre-recorded or live music; if relevant and feasible, accompany the music with instructions (e.g., guide breathing to relax the body, focus on different parts of the body, imaging of peaceful places).

•	After the listening experience, invite participants to share any changes in their levels of tension or emotions as observed while listening.

5.2.4.	Music listening with a focus on emotional expression

•	Invite participants to choose a piece that represents a certain event/feeling/challenge OR choose by yourself a piece to represent the group's main theme/challenge or to represent a participant in the group.

•	Listen to the musical piece.

•	Encourage a discussion around the meaning of the musical piece to the presenter and to the group, allow the expression of different feelings that might arise and con-tribute your own observations as appropriate.

5.2.5.	Music listening with a focus on lyric analysis

•	Choose a musical piece OR invite the participants to choose, according to a specific theme/title (e.g., recovery).

•	If relevant, provide lyrics on paper or on screen, read the lyrics out loud and trans-late if needed.

•	Play the pre-recorded piece, guide the participants’ attention while listening to rel-evant elements (e.g., lyrics that resonate with them).

•	Lead discussion (e.g., focusing on themes discussed in the group, such as motivation to recovery).

### 4. SOPs for monitoring long-term outcomes and follow-up visits

**4.1 Strategy for monitoring long-term outcomes in FALCO**

Objective

To ensure high-quality long-term clinical outcomes for the clinical trial, including primary and secondary clinical outcomes.

Overview

- In line with the SOP, all sites will be recommended to collect data in paper-and-pencil format and then enter it into REDCap following the four-eyes procedure. That solution allows us to compare the paper versions with the REDCap records to ensure data quality.
- For each site, the designated contact person will be asked to share the scans for the selected participants’ records in order to perform the data quality monitoring.
- The data monitoring will follow the four-eyes procedure, joining auditory and visual input.
- As the data monitoring procedure is typically performed in person, if the online setting is not efficient, two people will separately perform the data quality check and compare results.
- In total, every 10^th^ participant record will be monitored. For the first monitoring period (April 2026), it will be every 5^th^ participant record. For the next monitoring periods, it will be every 20^th^ participant record.
- If there are no paper records available, the data monitoring will include checking if the variability of the records is within the expected range and whether there is a systematic error across sites.

Data incident detection

- If an error/incident is detected, the following two participant records will be checked to investigate whether the error might be systematic. The site’s contact person will be notified of data discrepancies and incidents.
- If across the data monitoring, three data incidents are detected, the project PI will be notified.
- Efforts will be targeted at explaining the reason behind the data incidents. The discrepancies between the paper and the REDCap versions, if identified as data-entry errors, will result in reentry.

**4.2 Standard operating procedures for follow-ups in FALCO**

*Follow-up time windows*

Based on the current experience with the target population, which is particularly vulnerable and often difficult to reach, we propose the following time windows:

For the 3months: +1 month, -1 week

For 12 months: ±1 month

For 24 months: ±1 month

Moreover, if a participant misses a follow-up but has not officially withdrawn from the study, we still try to enroll them in the next follow-up.

*Contact attempts*

Regarding the duration and frequency of contact attempts, we propose using diversified contact strategies^1^, including a sequential mixed-mode design (e.g., SMS, phone, and email).

We suggest that diversified contact attempts should be made at least once per week over a period of up to two months, allowing for flexibility and increased likelihood of reaching participants.

If such data is available: contact relatives or close friends (as per protocol) or the clinical site that is in contact with the participant.

*Reminder: Retention strategies*

Several strategies should be implemented to ensure the retention of participants. These are summarized in the document “Retention strategies in FALCO.docx”. The document summarizes strategies pertaining to a) study design and participant requirements, b) staff requirements, and c) technical strategies that can be implemented to reduce attrition of participants over time.

### 5. Technical specifications of hair biomarkers

Three strands of hair as thick as a matchstick will be collected from the occiput as close to the scalp as possible. The 3 cm most proximal to the scalp will be analysed. The levels of substance use will be quantitatively determined by liquid chromatography-tandem mass spectrometry (LC-MS/MS) using a protocol deriving the maximum amount of high-quality valid information from the smallest amount of hair^2^. At each follow-up, the individual nominal change of the hair concentration of each substance will be determined. Additionally, for each substance, the change from baseline and follow-up will be scored into the following categories: recovery (min 75% decrease or below threshold^3^), remission (min 50% decrease), partial remission (min 25% decrease), no change (<25% decrease). The application of a brief hair quality questionnaire ensures the assessment of potential confounders of hair analyses (e.g., hair colour, bleaching, sun exposure, sports intensity, etc.). FALCO hair samples will be assayed for ≈100 substances, including:

1. Illegal substances and/or their metabolites such as THC and cannabinol (THC metabolite), cocaine and metabolites, MDMA and metabolites, amphetamines, opiates (e.g., heroin), and hallucinogens (e.g., ketamine, 2C-B).
2. Alcohol use intensity as measured with the marker ethylglucuronide (EtG)^4^.
3. Medications such as stimulants, antidepressants, antipsychotics, benzodiazepines, z-drugs, cough medication (codeine and dextromethorphan), non-opioid (e.g., diclofenac, paracetamol), and opioid pain killers (e.g., morphine, tramadol, tilidine, fentanyl, oxycodone, buprenorphine, etc.). Concentrations of opiates/opioids will be standardised according to morphine-equivalents.

### 6. Technical specifications of blood biomarkers

The sensitivity of NfL to substance-induced neurotoxicity has been initially shown in animal and in-vitro models using cocaine, ketamine, and opiates. In several clinical studies, SUDs involving cocaine, ketamine, alcohol, or polysubstance use were associated with elevated NfL levels^5^. GFAP has scarcely been studied in SUD yet, but preliminary data from the Quednow group at UZH show promising results of a similar sensitivity of GFAP for SUD compared to NfL. Thus, NfL and GFAP will be used as potential functional biomarkers of individual substance use burden of the brain. NfL, GFAP and p-Tau analysis: Plasma will be collected in K2EDTA tubes. The samples will be centrifuged with 2000 * g for 15 minutes at 4°C. Storage temperature will be -80°C. NfL, GFAP and p-Tau concentrations will be measured in triplicates using sim-plex assays (ProteinSimple, CA, USA) on Ella microfluidic system (BioTechne, MN, USA). To minimize the possibility of technical issues or batch effects, all samples will be analyzed together at the end of the study.

### 7. Technical specifications of structural brain imaging

Morphometric analysis (Magnetic resonance imaging (MRI)Voxel Based Morphometry, VBM): This analysis permits statistical estimation of whole-brain voxel-level local changes in volume. It will provide evidence of potential regional structural changes in grey matter volume or density observed due to MT [VBM will be implemented using the Cat12 toolbox (https://neuro-jena.github.io/cat/index.html#VBM) implemented in SPM12 (http://www.fil.ion.ucl.ac.uk/spm/)].

Cortical folding and cortical thickness: Geometrical representations for the cortical surfaces of each patient will also be reconstructed and compared based on 3D/T1 images. Specifically, cortical folding indices will be calculated by using BrainVisa database (http://brainvisa.info/), while measurements of cortical thickness, cortical surface area and subcortical volume will be performed using the FreeSurfer 7.1 software package (<http://surfer.nmr.harvard.edu>) (based on the following atlas: Destrieux - 74 bilateral cortical regions; Harvard-Oxford subcortical atlases, 11 bilateral regions). Concordance will be provided by comparing these results using also the Desikan-Killiany atlas.

Diffusion Tensor Imaging (MRI-DTI) analysis: DTI is unique in providing precise indices related to tissue white-matter integrity and microstructure reorganisation (i.e., Fractional anisotropy (FA) maps, or Radial diffusivity (RD), among others). A change in any of these indices can be associated with the physiological status of brain white structural connectivity and brain plasticity. We will use DW_MRI images to calculate FA and radial diffusivity maps by using the diffusion toolbox https://fsl.fmrib.ox.ac.uk/fsl). Additionally, two kinds of normalisation approaches will be assessed on these maps (to improve the objectivity and interpretability): (i) maps will be normalised using an optimised-protocol developed for DTI studies before voxel-wise statistical analysis and (ii) using a track-skeleton alignment projection in which all FA maps are projected, before applying voxel-wise cross-subject statistics.

DTI fiber tracking: Tracking and reconstruction of white-matter tracks allow to study the larger scale patterns of connectivity and differences in the major association paths in the brain. We will improve tractography by using advanced high-angular resolution diffusion imaging approaches which can capture multiple fibers directions within the same imaging voxel. Both deterministic and probabilistic tractography will be applied using both manual reconstructions as automatic tractography algorithms. We will focus on potential white-matter cortical-subcortical changes due to the potential effects of MT and related to reward-related and emotion regulation changes. Two white-matter connectivity pathways will be targeted: (i) potential pathways mediating the association between the ventromedial prefrontal and amygdala, associated with emotion regulation and (ii) the accumbofrontal track responsible for the connectivity between the orbitofrontal cortex and the ventral striatum, which has also been associated to hedonic musical responses ^6^.

### 8. Technical specifications of immediate outcomes

Comparing AMG and MLG to psychosocial interventions provided as part of TAU (Fig. SM7.1), this partly non-randomised embedded study aims to bridge between results of previous RCTs showing immediate effects^7^ to the longer-term outcomes addressed in this trial. As comparisons of immediate outcomes will be partly quasi-experimental (randomised to AMG or MLG, but non-randomised to other interventions), covariate adjustment will be important. The partly non-randomised design of this substudy and its statistical analyses will be described in a separate publication.

Motivation for change will be assessed with the Importance, Confidence, Readiness Ruler (ICR). This is a patient-reported scale to assess overall motivation to change, including readiness to change^8^. Three items are rated on a 10-point Likert scale with total scores ranging from 0-30, and a higher score indicating higher motivation to change. The scale has been used in previous MT trials^7^. Craving will be assessed using The Craving Scale^9^. Social connectedness with the members of the group session will be assessed using the Inclusion of Other in the Self Scale^10^. This is a patient-reported single-item visual scale of closeness and has been previously used in prior MT trials^11^. Scores range from 1-7, with higher scores indicating higher social connectedness.

**Figure SM7.1. Design of the FALCO sub-study on immediate outcomes**

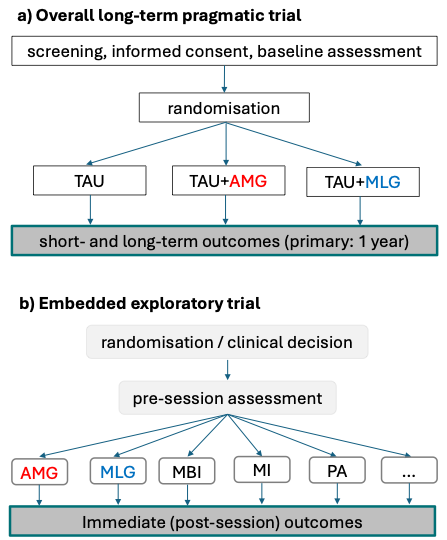

*Note*. AMG – active music group; MBI – mindfulness-based intervention; MI – motivational interviewing; MLG – music listening group; PA – physical activity; TAU – treatment as usual

### 9. Technical specifications of machine learning analyses

Supervised approach: The statistical analysis will be complemented by an interpretable ML-based approach for multivariable statistical analysis^12^. In contrast to in-sample inference, our ML analysis focuses on predictive accuracy, achieved through out-of-sample inference. This process involves generalising findings to new, unseen data using models that are flexible, complex, high-dimensional, yet remain robust and credible^12–15^. We will train the ML models to predict primary and secondary outcome measures from demographic, clinical, behavioural, and other relevant measures. Following the established suggestions regarding sample sizes and considering that a minimum of 10 samples per degree of freedom (predictor) are required, the current clinical trial with 450 participants will allow for including 45 predictor variables in the ML analysis^16,17^. For the supervised multivariate ML approach, we train Gradient Boosted Decision Tree (GBDT)^18^ models to perform the actual modelling task because this method is computationally efficient and highly accurate. GBDT models can inherently model multivariate associations and interactions between features and outcomes that go beyond one constant factor (nonlinear associations). In addition, GBDT models are robust to multicollinearity and outliers in the data due to sophisticated regularisation strategies^18^. For evaluation, post-hoc model fit is a problematic measure, as complex models will always achieve a very high model fit due to overfitting the data. A favourable alternative is out-of-sample prediction performance. Here, a model is evaluated in regard to its prediction performance on a new data set that was not present during training. This will be done via nested cross-validation (CV)^19^. CV implements repeated splitting of the data into training and test sets, thereby a group-controlled shuffle-split scheme will be applied in the main (outer) CV loop to deal with potential clusters. This scheme will allow for different types of grouping and stratification and, hence, for different levels of generalisation. In each iteration of the main CV loop, the respective training set will be used for data scaling (standardisation) and model complexity tuning. Model complexity tuning will be performed in a nested (inner) CV procedure using a random search scheme to find the best-performing parameters. Subsequently, the model will be tested on the corresponding test set of the main CV loop. Regression performance will be measured by the prediction R² and the mean absolute error (MAE)^20^. The prediction R² will be smaller than the R² values of conventional statistical models because the prediction R² measures the prediction performance for unknown data rather than the post-hoc model fit^20^. The MAE reflects the average error made on each prediction and is not normalised. Classification performance will be measured using weighted classification accuracy^20^. Here, a class will be up-weighted so that the total contribution of each class is equal. Subsequently, we will use SHapley Additive exPlanations^46^ to understand which features are associated with outcome^21^. SHAP is an interpretable ML method based on Shapley values that measures the contributions of each individual feature to a model's predictions, including interaction effects. Pooling these contributions across many predictions will allow for a comprehensive analysis of the influence of individual features on the prediction^22^. Subsequently, a modified t-test will assess the statistical significance of differences between the means of the prediction R² metric, prediction accuracy, and feature influence with their respective counterparts obtained under the simulated null hypothesis (shuffled data labels). The t-test will be modified to account for sample dependence due to CV. T-test results will be Bonferroni-corrected for multiple comparisons. This analysis procedure is inspired by Qiu et al.^21^ and is performed in Python v3.11 using the scikit-learn library v1.4^23^. Some scripts are available on Github ([https://github.com/univiemops/iml](https://eur04.safelinks.protection.outlook.com/?url=https%3A%2F%2Fgithub.com%2Funiviemops%2Fiml&data=05%7C02%7Casav%40norceresearch.no%7Cc75ee5bccb714a45393c08de2d9b968f%7C70a6eba4967145d2b83e432e06502242%7C0%7C0%7C638998343440042766%7CUnknown%7CTWFpbGZsb3d8eyJFbXB0eU1hcGkiOnRydWUsIlYiOiIwLjAuMDAwMCIsIlAiOiJXaW4zMiIsIkFOIjoiTWFpbCIsIldUIjoyfQ%3D%3D%7C0%7C%7C%7C&sdata=3NIGzuBFKR3%2FcvKREMx0XffX2cugSB1bP2HzgY4qapM%3D&reserved=0)).

Unsupervised ML approach: We will use a data-driven, unsupervised multivariate ML approach as a complimentary test of predictor variables to outcome measure relationships. This approach will use t-distributed Stochastic Neighbour Embedding (t-SNE) for dimensionality reduction, hierarchical density-based spatial clustering of applications within noise (HDBSCAN)^12^ for clustering, and finally, we will relate the found clusters to the outcome measures^24,25^. While the supervised learning approach potentially will be the more powerful method for finding subtle relationships, the unsupervised approach will be more robust to unreliable measures and differences in measurement instruments of outcomes. Furthermore, this approach is resistant to overfitting because the outcome measures are not used in dimensionality reduction and clustering of the predictor variables. It is a nonlinear dimensionality reduction method for embedding high-dimensional data in a low-dimensional space of two or three dimensions^24^. Specifically, t-SNE models each high-dimensional data point with a two- or three-dimensional point so that similar objects are modelled by nearby points and dissimilar objects by distant points with high probability. This means participants with similar characteristics will group together in a low-dimensional space. HDBSCAN will subsequently be performed on the low dimensional t-SNE embeddings of the data. The HDBSCAN algorithm can be considered as an extension of DBSCAN. Specifically, DBSCAN assumes that the clustering criterion (i.e., the density requirement) is globally homogeneous. As a result, DBSCAN may have difficulty successfully detecting clusters with different densities. HDBSCAN relaxes this assumption and explores all possible density scales by building an alternative representation of the clustering problem, thereby automatically selecting the number of clusters in a data-driven way. Finally, we will look for relationships between obtained clusters and outcomes, using mutual information as a measure of agreement^26^.

### 10. Ten-year outcomes

Patients’ participation in the study ends after two years. From electronic health records and other public records that are unaffected by loss of contact, employment, hospitalisation, criminal record and survival will be monitored until 10 years after randomisation. Registry data will be analysed as binary or time-to-event data. While included in consent forms, the actual collection and analysis of these very long-term outcomes is beyond the scope of the current funded project and will require separate funding.
